# Simple decision rules to predict local surges in COVID-19 hospitalizations during the winter and spring of 2022

**DOI:** 10.1101/2021.12.13.21267657

**Authors:** Reza Yaesoubi, Shiying You, Qin Xi, Nicolas A. Menzies, Ashleigh Tuite, Yonatan H. Grad, Joshua A. Salomon

## Abstract

Low rates of vaccination, emergence of novel variants of SARS-CoV-2, and increasing transmission relating to seasonal changes leave many U.S. communities at risk for surges of COVID-19 during the winter and spring of 2022 that might strain hospital capacity, as in previous waves. The trajectories of COVID-19 hospitalizations during this period are expected to differ across communities depending on their age distributions, vaccination coverage, cumulative incidence, and adoption of risk mitigating behaviors. Yet, existing predictive models of COVID-19 hospitalizations are almost exclusively focused on national- and state-level predictions. This leaves local policymakers in urgent need of tools that can provide early warnings about the possibility that COVID-19 hospitalizations may rise to levels that exceed local capacity. In this work, we develop simple decision rules to predict whether COVID-19 hospitalization will exceed the local hospitalization capacity within a 4- or 8-week period if no additional mitigating strategies are implemented during this time. These decision rules use real-time data related to hospital occupancy and new hospitalizations associated with COVID-19, and when available, genomic surveillance of SARS-CoV-2. We showed that these decision rules present reasonable accuracy, sensitivity, and specificity (all ≥80%) in predicting local surges in hospitalizations under numerous simulated scenarios, which capture substantial uncertainties over the future trajectories of COVID-19 during the winter and spring of 2022. Our proposed decision rules are simple, visual, and straightforward to use in practice by local decision makers without the need to perform numerical computations.

**Significance Statement:** In many U.S. communities, the risk of exceeding local healthcare capacity during the winter and spring of 2022 remains substantial since COVID-19 hospitalizations may rise due to seasonal changes, low vaccination coverage, and the emergence of new variants of SARS-CoV-2, such as the omicron variant. Here, we provide simple and easy-to-communicate decision rules to predict whether local hospital occupancy is expected to exceed capacity within a 4- or 8-week period if no additional mitigating measures are implemented. These decision rules can serve as an alert system for local policymakers to respond proactively to mitigate future surges in the COVID-19 hospitalization and minimize risk of overwhelming local healthcare capacity.

## Introduction

Many communities are at risk of surging COVID-19 hospitalizations during the winter and spring of 2022 due to low rates of vaccination, emergence of novel variants of SARS-CoV-2, and seasonal changes in transmission (1). Understanding the likely trajectory of the pandemic and its implications for demands on the healthcare system are important for policymakers aiming to prepare for and possibly prevent surges that result in hospital demand that exceeds capacity (2). Since the beginning of the pandemic, substantial efforts have been invested in developing models to predict the trajectories of cases, hospitalizations, and deaths associated with COVID-19 (e.g., COVID-19 Forecast Hub (3) or the IHME COVID-19 Forecasting Model (4)). While the spread of SARS-CoV-2 and hospitalizations due to COVID-19 vary substantially across different geographic regions (as influenced by a population’s characteristics, local policies, and adoption of risk-mitigating behaviors), these models typically focus on predictions at national or state levels. This leaves local policymakers in urgent need of tools that can signal when risks are high for overwhelming local hospital capacity with COVID-19 cases in the absence of additional mitigation measures.

Local trajectories of COVID-19 during the winter and spring months will be impacted by various factors, including the proportion of the population with infection-or vaccine-induced immunity, the duration of infection-and vaccine-induced immunity, uptake and effectiveness of vaccine boosters, the transmissibility and virulence of novel variants (such as the omicron variant) that may continue to emerge and spread, the effectiveness of vaccines against prevalent strains including novel variants, and population behavior and adherence to mitigating strategies (1, 2, 5–8). True values of these pandemic parameters and state variables are either unobservable or can only be estimated with a high level of uncertainty, which further challenges our ability to predict local trajectories of COVID-19 pandemic (2).

Data from hospital occupancy censuses, rate of new COVID-19 hospital admissions, and vaccination coverage are often available to monitor the local spread of SARS-CoV-2 and trends in COVID-19 hospitalizations. To enable local policymakers to translate the data from these surveillance systems into timely decisions, this study aims to identify simple and easy-to-communicate decision rules to provide early warnings when a pre-specified threshold of hospital capacity is likely to be exceeded within a 4- or 8-week period. We evaluate the predictive accuracy of these simple decision rule using simulated trajectories from a model that incorporates complexities and uncertainties regarding the biology of SARS-CoV-2 and factors driving local trajectories of COVID-19 during the winter and spring of 2022.

## Methods

### Overview

Multiple indicators are collected through surveillance systems to monitor and predict local trends in COVID-19 hospitalizations (Table 1). We use these indicators (which we will refer to as ‘features’) to summarize the information from each surveillance system into a number of predictors (e.g., the average change in the number of hospitalizations during the past 4 weeks or the number of individuals vaccinated thus far). Our goal is to develop decision trees to predict if the local hospitalization capacity will be surpassed within 4 or 8 weeks based on the values of features defined in Table 1. Decision tree models provide simple, visual, and explicit decision rules to predict the outcomes of interest, which makes them straightforward to use in practice (9, 10).

**Table 1:** Observations available through surveillance systems to predict the local trend in COVID-19 hospitalizations during the winter and spring of 2022.

| Surveillance | Features used for prediction |
| --- | --- |
| Rate of hospital occupancy due to COVID-19 | - Current value |
| Weekly rate of new hospital admissions due to COVID-19 | - Average over the past 2 weeks<br>- Average change during the past 4 weeks |
| Vaccination coverage | - Cumulative value |
| Prevalence of novel variant among new infections | - Average over the past 2 weeks<br>- Average change during the past 4 weeks |

Predictive models are usually trained on historical data. If the process that generates data does not substantially change over time, models trained on historical data could provide accurate predictions in the future. In the context of COVID-19 pandemic, however, the assumption of a stationary data-generating process does not necessarily hold. The factors impacting the observations related to COVID-19 (e.g., the timing and the effectiveness of mitigating strategies, the characteristics of novel variants, and the coverage of vaccination among different age groups) will most likely continue to change over the coming months. The types and the effectiveness of mitigating strategies during the near future could be markedly different than those employed in the past, novel variants such as omicron may gain hold over different time courses in different locations, and their characteristics in terms of transmissibility and virulence will be highly uncertain during their initial seeding, and vaccination coverage trends are also uncertain and contingent. Hence, to develop decision trees that are robust against changes in the data generating process and future uncertainties, we use epidemic trajectories simulated by a model of SARS-CoV-2 transmission in the U.S. between March 1, 2020 and June 1, 2022 to train and evaluate our decision trees.

This simulation model is structured to incorporate factors, and the associated uncertainties, that impact the local size of COVID-19 hospitalizations during the winter and spring of 2022 (Table 2). To build the datasets needed for our purpose, we use a set of simulated trajectories that satisfy specific epidemiological conditions during March 1, 2020 and November 30, 2021. These conditions, which relate to the historical rate of hospitalization (overall and by age), age-distribution of hospitalizations, the prevalence of population with immunity against SARS-CoV-2, the spread of the delta variant, and the rate of vaccination (overall and by age), ensures that the selected trajectories are consistent with past trajectories of COVID-19. We then project these selected trajectories onto the period of the winter and spring of 2022 to build the datasets needed to train and evaluate our decision trees. The details of this simulation model and the process to select trajectories are provided below.

**Table 2:**
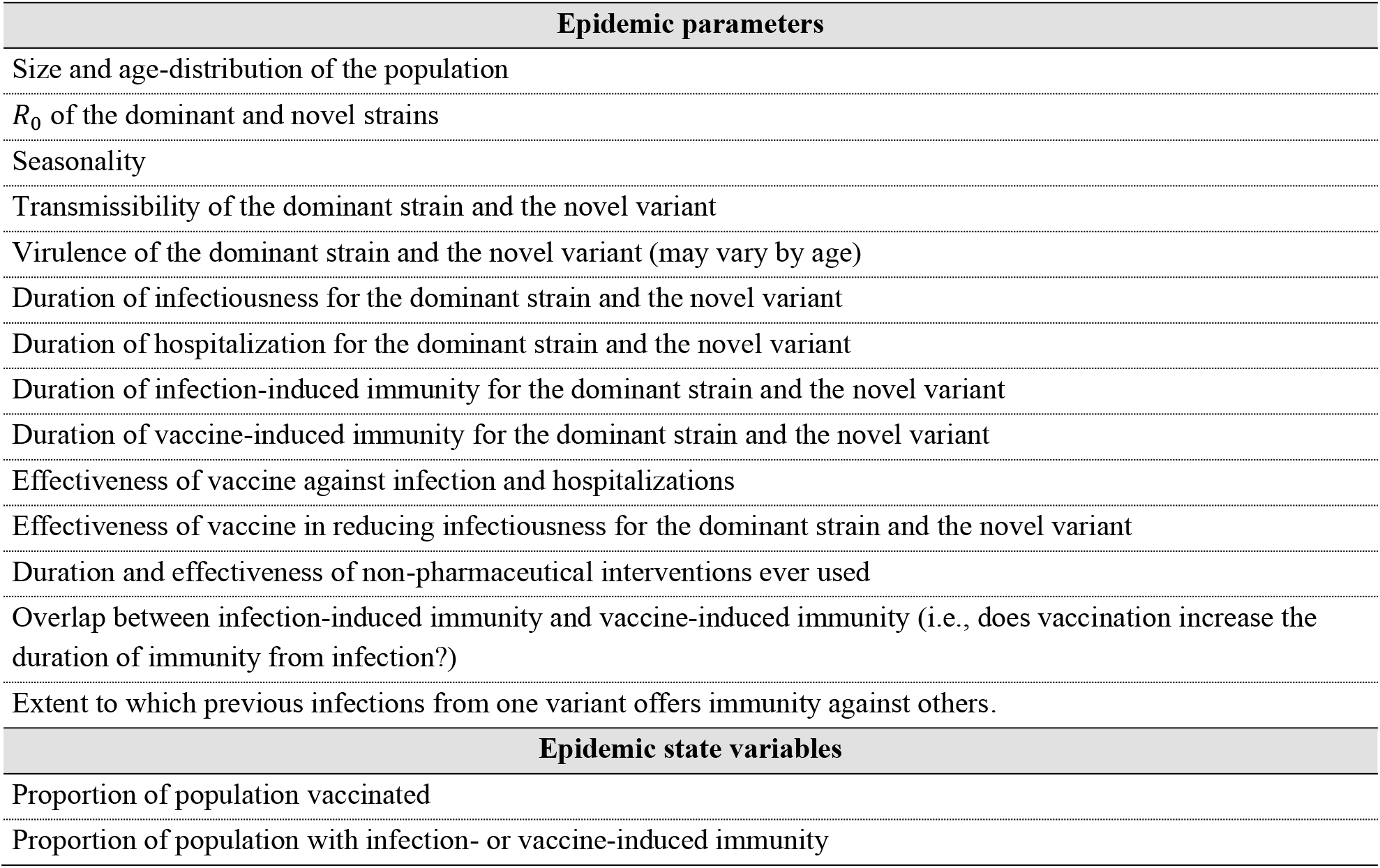
Factors that could influence the local trajectory of COVID-19 hospitalizations during the winter and spring of 2022 (1, 2, 5–8).

| Epidemic parameters |
| --- |
| Size and age-distribution of the population |
| $R_0$ of the dominant and novel strains |
| Seasonality |
| Transmissibility of the dominant strain and the novel variant |
| Virulence of the dominant strain and the novel variant (may vary by age) |
| Duration of infectiousness for the dominant strain and the novel variant |
| Duration of hospitalization for the dominant strain and the novel variant |
| Duration of infection-induced immunity for the dominant strain and the novel variant |
| Duration of vaccine-induced immunity for the dominant strain and the novel variant |
| Effectiveness of vaccine against infection and hospitalizations |
| Effectiveness of vaccine in reducing infectiousness for the dominant strain and the novel variant |
| Duration and effectiveness of non-pharmaceutical interventions ever used |
| Overlap between infection-induced immunity and vaccine-induced immunity (i.e., does vaccination increase the duration of immunity from infection?) |
| Extent to which previous infections from one variant offers immunity against others. |
| Epidemic state variables |
| Proportion of population vaccinated |
| Proportion of population with infection- or vaccine-induced immunity |

### A simulation model of SARS-CoV-2 transmission

We developed a stochastic, age-structured model that describes the transmission of three main variants of SARS-CoV-2 between March 1, 2020 and June 1, 2022 (Fig. 1). The variants represent the ancestral strain of SARS-CoV-2 that dominated during 2020, the delta variant that began spreading in the Spring of 2021, and a novel variant, such as omicron, that could overtake delta (11, 12). The model projects the weekly incidence of cases, hospitalization, and deaths due to COVID-19 among age groups 0-4, 5-12, 13-17, 18-29, 30-49, 50-64, 65-74, and 75+ in communities with population between 250,000 and 1,250,000. The mixing patterns between age groups are modeled using the age-specific contact rates estimated for the U.S. population (13) (see §S2.3 of the Supplement).

**Fig. 1:**
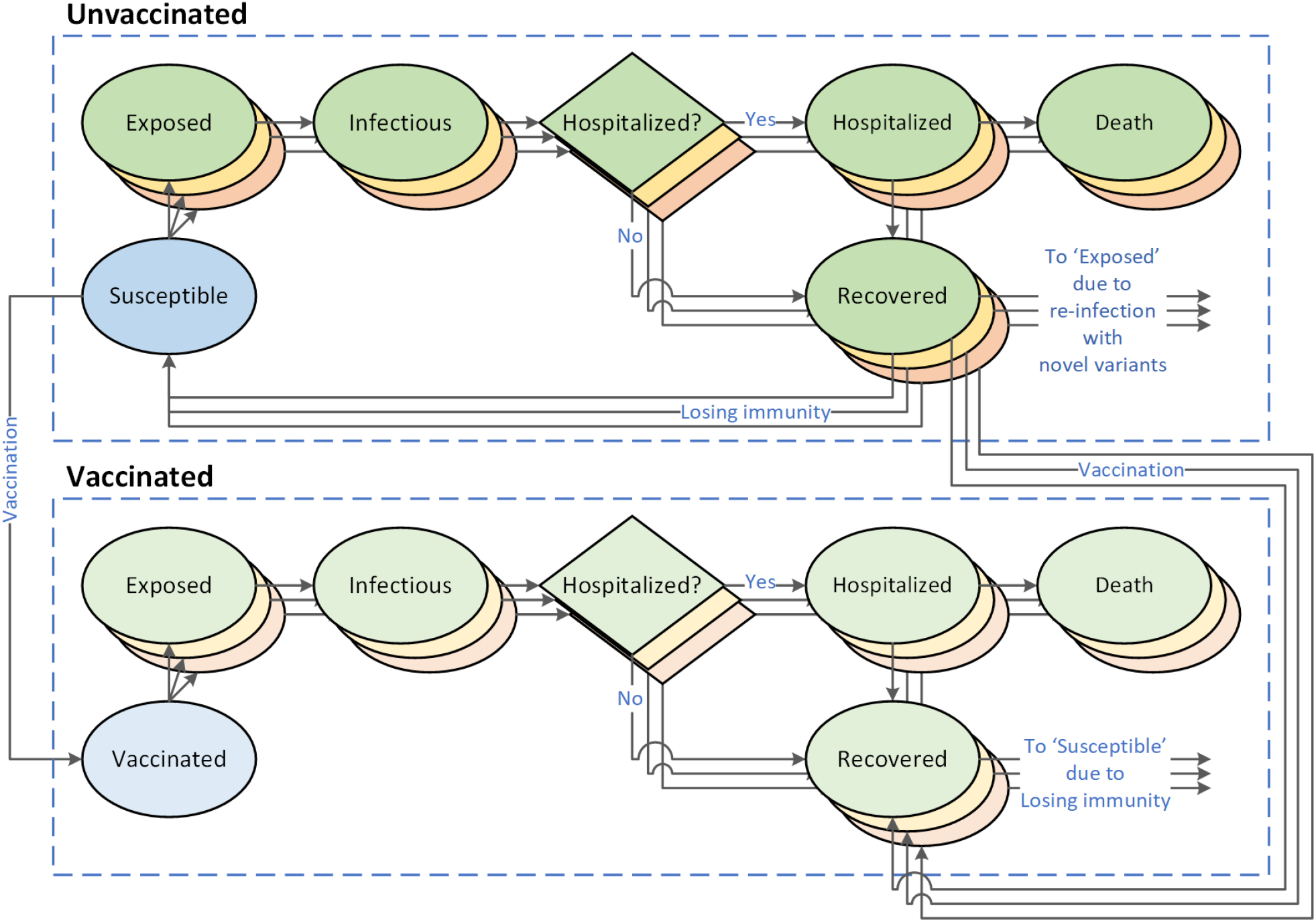
A stochastic, age-structured model of SARS-COV-2 transmission with three strains and two vaccination status. The green, yellow, and red compartments represent, respectively, the ancestral strain of SARS-CoV-2, the delta variant, and a novel variant of SARS-CoV-2 (such as the omicron variant) that might emerge and spread during the winter and spring of 2022.

As the model attempts to describe the transmission of SARS-CoV-2 at the local level, we allow for a continuous importation of cases from neighboring communities. An imported case could be infected with the novel variant with a probability that begins to increase around December 2021 according to a sigmoid function (Fig. S1). Given the uncertainty in the timing for the introduction of the novel variant, we allowed the magnitude of this probability and the rate at which it increases over time to vary across simulated trajectories (Fig. S1 and Table S3).

We assumed that compared to the current dominant strain, the novel variant could be more transmissible (up to twice) (14, 15), could lead to milder or more severe disease (up to 200% increased or 100% decreased probability of hospitalization) (12, 16, 17), could cause a shorter or longer duration of infectiousness (up to 100% increase or decrease), and could evade immunity conferred by previous infection or vaccination (Table S3 and Table S4).

We assume that vaccination begins in December 2020 at an age-specific rate that gradually decreases over time (Fig. S2 and Table S5). For the ancestral strain of SARS-CoV-2, vaccine provides 85%-100% effectiveness against hospitalization and reduces the duration of infectiousness by 25%-75% (18–20). Our model does not differentiate vaccinated individuals based on the type of vaccine or the number of vaccine doses they have received; therefore, we consider an individual “vaccinated” when they can be assumed to have reached the level of immunity described above. We assume that vaccine-induced immunity wanes (within 0.5-2.5 year) leading to the vaccinated individual becoming susceptible (Fig. 1).

With respect to the novel variant, vaccinated individuals are assumed to have partial immunity to infection (up to 100%), and if infected, experience a shorter duration of infectiousness by 25%-50% (20) and are 50-100% less likely to require hospitalization (Table S5). We also assume that vaccination increases the duration of infection-induced immunity by up to 50% for both the dominant strain and novel variant (Table S5).

To model the effect of control measures and population adherence to public health recommendations across different communities and since the beginning of the pandemic, we assume that control measures went into effect whenever the rate of hospital occupancy due to COVID-19 exceeded the threshold *T*_1_ and were lifted whenever this rate dropped below the second threshold *T*_2_ (21). We further assumed that the intensity and the effectiveness of control measures in reducing the effective reproductive number increase with the rate of hospital occupancy according to some sigmoid function (Fig. S3). To account for the variation in timing and effectiveness of control measure across different communities, we allowed the thresholds *T*_1_ and *T*_1_, and the function that models the effectiveness of control measure to vary across simulated trajectories and to be determined by random draws from appropriate probability distributions (see §S2.4 of Supplement).

### Modeling errors in surveillance estimates

The estimates provided by certain surveillance systems are subject to error due to limited or unrepresentative samples. Among the surveillance systems of Table 1, we assume that the rate of hospital occupancy, the weekly rate of new hospitalizations, and vaccination coverage can be observed with no error in each community. The accuracy of estimates for the prevalence of a novel variant among the new infections depends on the number of samples collected and tested. To account for this sampling error, we use the following approach. Let *y*_*t*_ be the true value of the prevalence of a novel variant among infections in week *t* of the pandemic. We assume that *y*_*t*_ can be observed (denotated by *ŷ*_*t*_) with some error (*ε*_*t*_) and a delay of one week (22):

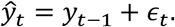

Here, we assume that *ε*_*t*_ follows a normal distribution with mean 0 and standard deviation 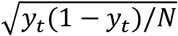 where *N* is the sample size of the survey. A higher value of *N* decreases the variance of the error *ε*_*t*_ leading to more accurate estimates. The exact number of samples that are tested for novel variants per week is unclear (22); therefore, we assumed that enough samples are collected to estimate the prevalence of 1% with the 95% confidence interval of (0.5-1.5%). This requires a sample size of *N* = 1521 per week.

### Selection of simulated trajectories to develop and evaluate decision tree models

To ensure that the trajectories simulated by our model are consistent with the community-level spread of SARS-CoV-2 in the U.S., we only considered trajectories that satisfied specific conditions related to the rate of hospitalization (overall and by age), age-distribution of hospitalizations, prevalence of population with immunity against SARS-CoV-2, the prevalence of the delta variant among new infections, and the rate of vaccination (overall and by age). Additional details are provided in §S3 of the Supplement. This selection approach seeks to identify simulated trajectories that are consistent with the actual state-level trajectories of COVID-19 in the U.S. but also describe the additional variation and uncertainties in the trajectories of COVID-19 across more granular geographic regions. Here, each selected trajectory represents a community with unique values for the population characteristics (e.g., size and age distribution), effect of mitigating strategies, and other factors that determine the trajectory of COVID-19 hospitalizations during the winter and spring of 2022 (as described in Table 2).

### Decision tree models

We considered two decision tree models which differed based on the surveillance systems they use to predict whether the hospital occupancy due to COVID-19 would surpass the hospitalization capacity in the next 4 or 8 weeks over the winter and spring months. Decision Tree A uses the information related to the current hospital occupancy, the weekly rate of new hospitalizations, and the vaccination coverage at the time of prediction. Decision Tree B augments Decision Tree A by assuming access to the percentage of weekly incidence due to novel variant, available through genomic surveillance of SARS-Co-V-2 (23).

We trained each model to predict whether hospital occupancy due to COVID-19 exceeds the hospitalization capacity of 15 per 100,000 population (24) within the next 4 or 8 weeks. We also established decision rules when hospitalization capacity is 10 or 20 per 100,000 population. To avoid overfitting, we used a minimal cost-complexity pruning approach (10), where we determined the complexity parameter using 10-fold cross-validation to maximize the model accuracy (defined as the fraction of correct predictions) (9). We used 2,000 simulated trajectories to train and optimize the parameters of each decision tree and used a separate set of 500 simulated trajectories to evaluate the final accuracy of each model. To build the datasets to develop and validate our predictive models, for each simulated trajectory, we recorded the values of features defined in Table 1 at weeks 0, 2, 4, …, 16, and 20 after the start of winter 2022. For each recording, the outcome of interest to predict was whether the hospital occupancy would surpass a prespecified threshold within 4 or 8 weeks.

In addition to the estimated accuracy of each decision tree model, we also report the model’s sensitivity (i.e., the probability of correctly predicting the event where hospitalization capacity will be surpassed within the projection period of 4 or 8 weeks), and specificity (i.e., the probability of correctly predicting the event where hospitalization capacity will not be surpassed within the projection periods). For each model, we estimated the accuracy, sensitivity, and specificity using a separate set of simulated trajectories not used to train these models.

### Sensitivity analyses

While we developed and validated our decision trees using a wide range of simulated trajectories, we also evaluated whether the accuracy of our predictive models persists under three extreme scenarios:

1. In our main analysis, we assumed that some forms of non-pharmaceutical measures (e.g., physical distancing and mask use recommendations) with varying degrees of effectiveness would remain in effect during winter and spring of 2022. Our first sensitivity analysis considers a scenario in which all non-pharmaceutical measures are removed (or the adherence to public health recommendations is minimal due to public fatigue) during this period (1).
2. In our main analysis, we assumed that a novel variant with uncertain degree of transmissibility and virulence emerges and spreads during winter and spring of 2022. Our second sensitivity analysis considers the scenario where no novel variant spreads during this period.
3. Finally, we trained our predictive models assuming that the prevalence of novel variant among new infections are estimated using the sample size of *N* = 1521 test per week (as described above, this was calculated based on the assumption that enough samples are collected to estimate the 1% prevalence of the novel variant with the 95% confidence interval of (0.5-1.5%)). Our third sensitivity analysis considered the scenario where a smaller number of samples (*N* = 250) are tested for infection with novel variant.

## Results

Fig. 2 demonstrates that our selected simulated trajectories to train our decision trees are consistent 1) with historical data on hospitalizations due to COVID-19, vaccination coverage in the U.S., and the spread of the delta variant; and 2) with the latest data regarding the state of the pandemic at the beginning of winter 2022 (i.e., week 91 in Fig. 2). The red regions in Fig. 2A–C represent the feasibility conditions for including a simulated trajectory to train and evaluate our predictive models with respect to the weekly rates of hospital occupancy, new hospitalizations, and the prevalence of population with immunity from infection (see §S3 for details). The age-specific rates of cumulative hospitalization and vaccination as well as the age-distribution of cumulative hospitalizations in our selected trajectories were also consistent with the reported data (Fig. S5), which demonstrate the ability of our model to properly describe the transmission of SARS-CoV-2 among different U.S. age groups. Fig. 2C–E show that our selected trajectories are representative of the state of the pandemic at the end of November 2021 as determined by the prevalence of population with immunity from infection (Fig. 2C), the rate of cumulative hospitalization (Fig. 2D), the prevalence of vaccinated individuals (Fig. 2E).

**Fig. 2:**
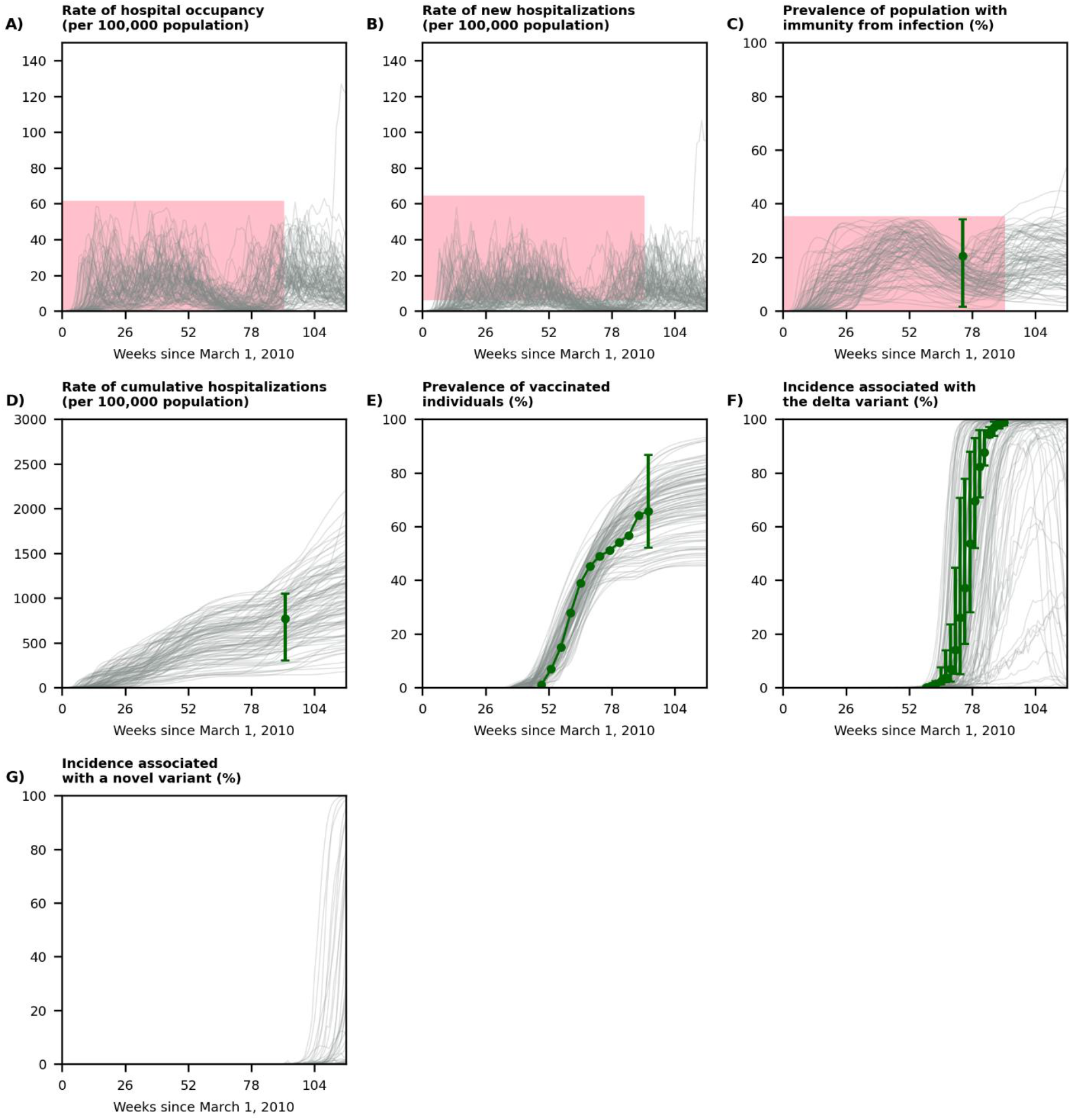
Displaying a random set of 100 trajectories simulated by our model (out of 1,000 simulated trajectories used to develop our decision trees). The week 91 marks the beginning of winter 2022. The red regions represent the feasibility conditions for including a simulated trajectory to train and evaluate the predictive models (see §S3 for details). The green dot in **panel C** is the prevalence of individuals with immunity againts SARS-CoV-2 and the interval represent the minimum and maximum prevalence in U.S. states as estimated by the CDC’s seroprevalence survey (26). The green dot in **panel D** is the cumulative hospitalization rate in the U.S. and the interval represents the minimum and maximum cumulative hospitalization rates observed in the surveillance sites of COVID-NET on November 27, 2021 (Table S7). The green dots in **panel E** represent the vaccination coverage provided by COVID data tracker, defined as the percentage of the population fully vaccinated (**Table S8**) and the interval represented the minimum and maximum vaccination coverage in all states (Table S9) on December 7, 2021. The green dots in **panel F** represent the prevalence of delta variant among new cases estimated by the CDC’s COVID Data Tracker; the intervals represent the minimum and the maximum values observed among 10 U.S. regions (Table S10). See Fig. S5 for the behavior of selected trajectories with respect to age-specific targets.

With respect to the spread of novel variants, Fig. 2F compares the proportion of weekly incidence associated with the delta variant in our selected trajectories with the estimated prevalence of the delta variant in the U.S. during April and August 2021. Fig. 2G displays the potential spread of a novel variant starting in the winter. For some trajectories, the spread of the novel variant was similar to that of the delta variant in the U.S., but for others, the spread was faster or slower depending on the characteristics of the novel variant.

There were substantial variations across our selected trajectories induced by various uncertain factors that influence the medium-term future trajectory of COVID-19 (Table 2). Among our selected trajectories, 92.3%, 82.1%, and 70.2% surpassed the hospitalization capacities of 10, 15, and 20 per 100,000 population during the winter and spring (Fig. 2A); the peak of hospital occupancy due to COVID-19 was in 95^th^ percentile range (4.8, 59.2) with mean 29.4 per 100,000 population (Fig. 2A); the peak of new hospital admission rate was in 95^th^ percentile range (4.2, 51.7) with mean 24.8 (Fig. 2B); and the prevalence of the population with immunity from infection varied between (0.8%, 50.8%) (Fig. 2C). Among these trajectories, by July 1, 2022 the rate of cumulative hospitalization since the beginning of the pandemic would be in 95^th^ percentile range (267.2, 1800.9) with mean 983.6 per 100,000 population (Fig. 2D), and the prevalence of vaccinated individuals would be in the 95^th^ percentile range (44.8, 90.5) with mean 68.7 (Fig. 2E). The prevalence of novel variant among new infections reached 5% among 15.6% of selected trajectories and reached 95% among 3.5% of the selected trajectories (Fig. 2F) during the winter and spring of 2022.

The final dataset we used to develop our decision tree models included 7000 records and the hospital capacity surpassed the thresholds of 10, 15, and 20 per 100,000 population within 8 weeks in 82.1%, 66.8%, and 51.4% of these simulations. The correlations between the features defined in Table 1 and the event that hospital capacity surpassed the above thresholds within 4 or 8 weeks were all significant (Table S11-Table S12).

Pruned Decision Trees A and B are shown in Fig. 3. Decision Tree A uses surveillance data related to hospital occupancy, the weekly rate of new hospitalizations, and the vaccination coverage to predict whether the hospital occupancy due to COVID-19 would surpass the threshold of 15 per 100,000 population within the next 8 weeks (see Fig. S9 in the Supplement for 4-week projections). Among the features used by this model, three identified as important after optimizing the parameters of the tree: 1) current hospital occupancy due to COVID-19 (per 100,000 population), 2) rate of weekly new COVID-19 hospitalizations averaged over past 2 weeks (per 100,000 population), and 3) change in weekly new COVID-19 hospitalizations over the past 4 weeks (per 100,000 population). Using the validation dataset, we estimated the sensitivity and specificity of this model at 0.936 and 0.833. This decision tree maintains its performance under extreme scenarios that we considered in our sensitivity analyses (Table 3).

**Table 3:** Performance of Decision Trees A and B (Fig. 3) evaluated using 500 simulated trajectories not used for training these models

|  | Accuracy | Sensitivity | Specificity |
| --- | --- | --- | --- |
| **Base scenario** |  |  |  |
| Decision Tree A | 0.866 | 0.936 | 0.833 |
| Decision Tree B | 0.870 | 0.851 | 0.879 |
| **If non-pharmaceutical measures are removed during the winter and spring of 2022** |  |  |  |
| Decision Tree A | 0.959 | 0.935 | 0.962 |
| Decision Tree B | 0.961 | 0.849 | 0.974 |
| **If no novel variant emerges during the winter and spring of 2022** |  |  |  |
| Decision Tree A | 0.887 | 0.944 | 0.858 |
| Decision Tree B | 0.876 | 0.821 | 0.905 |
| **Genomic surveillance with small sample size ($N=250$ tests per week)** |  |  |  |
| Decision Tree A | 0.866 | 0.936 | 0.833 |
| Decision Tree B | 0.870 | 0.851 | 0.879 |

**Fig. 3:**
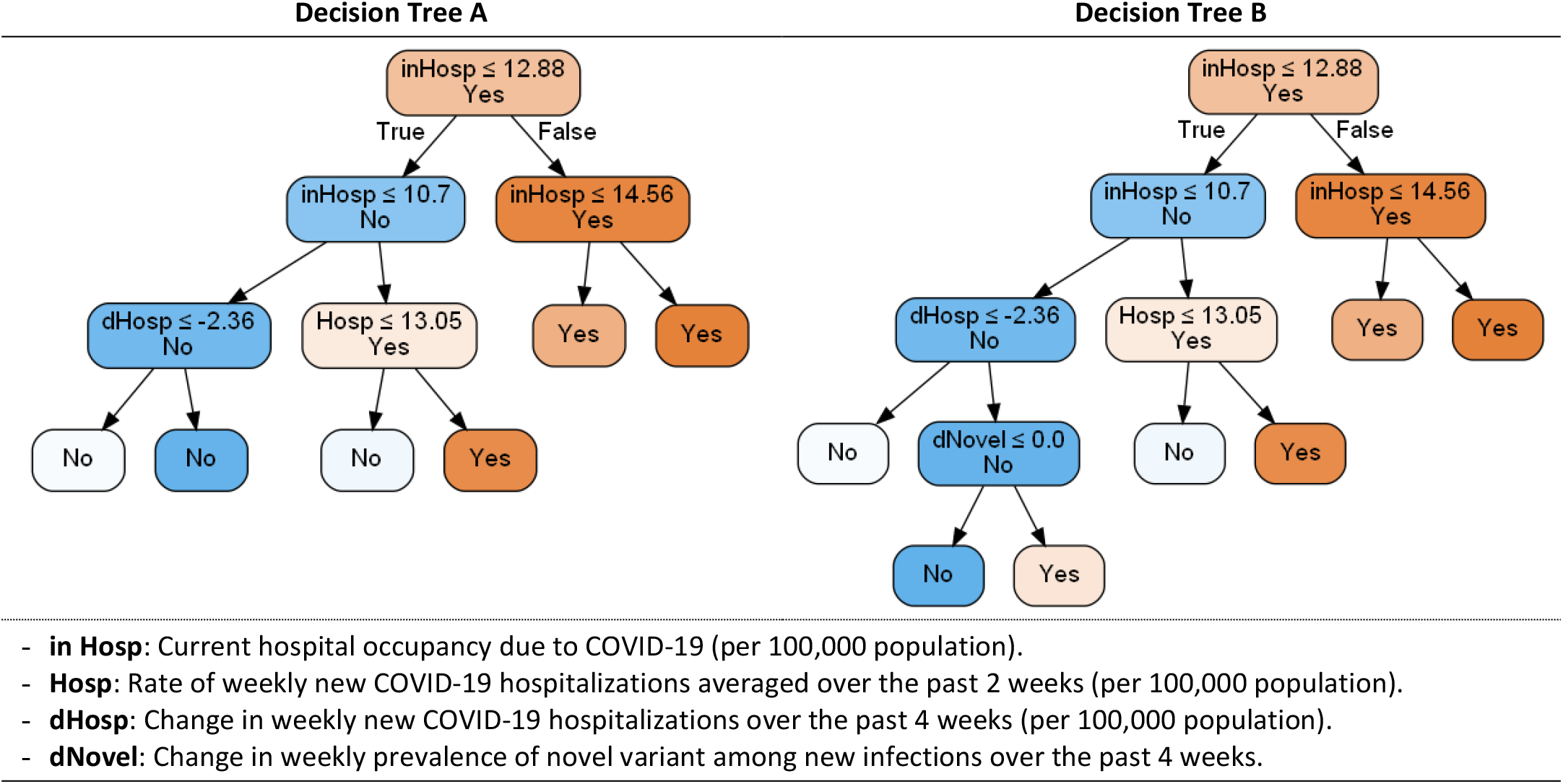
Decision Trees A and B to predict whether the hospital occupancy due to COVID-19 would surpass the threshold of 15 per 100,000 population within the next 8 weeks during the winter and spring of 2022. ‘Yes’ denotes the prediction that hospital occupancy will surpass the capacity and ‘No’ denotes the prediction that hospital occupancy will remain below the capacity. Between two descendent nodes, darker color indicates higher proportion of observations reaching the node. Decision trees for hospitalization capacity of 10 and 20 per 100,000 population are shown in Fig. S6-Fig. S7 and decision trees for 4-week predictions are shown in Fig. S8-Fig. S10.

In addition to the information available to Decision Tree A, Decision Tree B uses data from genomic surveillance systems to predict whether the hospital occupancy due to COVID-19 would surpass the threshold of 15 per 100,000 population within the next 8 weeks (see Fig. S9 in the Supplement for 4-week projections). The optimized Decision Tree B utilizes four features: change in weekly prevalence of novel variant among new infections over the past 4 weeks in addition to the three features identified as important by Decision Tree A. Using the validation dataset, we estimated the sensitivity and specificity of this model at 0.851 and 0.879. The performance of Decision Tree B also remains robust under extreme scenarios that we considered in our sensitivity analyses (Table 3).

The structure of the proposed decision trees (Fig. 3 and Fig. S6-Fig. S10) reveals that the signals in current hospital occupancy and weekly rate of new hospitalizations are strong enough to accurately predict short- and mid-term surges in hospitalizations despite the substantial uncertainty in factors that determine the local trajectories of COVID-19. The structures of our decision trees also suggest that the estimates of vaccination coverage do not contribute to the accuracy of predictions and the estimates for the prevalence of novel variant among new infections would slightly improve the 8-week predictions only if the hospital occupancy due to COVID-19 is relatively low. Once the rate of hospital occupancy reaches a certain threshold, the transmissibility and virulence of the novel variant is reflected in the hospitalization data; hence, the contributions of estimates for the prevalence of novel variant among new infections would be minimal (Table 3 and Table S13-Table S17 in the Supplement).

## Discussion

We sought to identify simple, easy-to-communicate decision rules that use surveillance data to alert local U.S. policymakers when hospitalizations due to COVID-19 during the winter and spring of 2022 are expected to surpass the local health care capacity within the next 4 or 8 weeks. To identify and evaluate these decisions rules, we used thousands of simulated trajectories representing communities with different characteristics that determine the burden of COVID-19, such as population size, age structure, vaccination uptake, effectiveness of mitigating strategies, and the population’s adherence to public health recommendations (Table 2). Our analysis suggests that decision rules that uses data on current hospital occupancy and the weekly rate of new hospitalizations due to COVID-19 could achieve a high level of sensitivity and specificity in predicting whether hospitalization capacity would be surpassed in the next 4 or 8 weeks. Access to the estimates for vaccination coverage or the prevalence of novel variant among new infections does not markedly improve the performance of these decision rules (Table 3 and Table S13-Table S17 in the Supplement).

There is substantial uncertainty in how the COVID-19 pandemic would impact local communities during the upcoming spring and winter. This is caused by uncertainties in factors such as the effect of seasonality in the transmission of SARS-CoV-2, the duration of infection- and vaccine-induced immunity, the transmissibility and virulence of novel variants such as omicron and others that may emerge, the vaccine effectiveness against the prevalent and novel variants, and the population’s adherence to public health recommendations during this period (Table 2). Using simulated trajectories distinct from those used to characterize our decision rules, we showed that the accuracy, sensitivity, and specificity of our proposed decision rules are robust to the substantial level of uncertainties surrounding the future of the COVID-19 pandemic at the local level. The performance of these decision rules maintains under extreme scenarios where all non-pharmaceutical interventions are lifted, no novel variant emerges and spreads, and capacity of genomic surveillance is substantially reduced (Table 3 and Table S13-Table S17 in the Supplement)).

Our study has a number of limitations. First, predicting the future trajectory of COVID-19 hospitalizations is challenged by various barriers, some of which are due to uncertainties in epidemic parameters and state variables (Table 2). Although our analysis accounts for these sources of uncertainties, predicting the local trajectories of COVID-19 are further challenged by the unpredictability of population’s behavior and policymakers’ responses to a slowing or speeding pandemic. To minimize the impact of these unpredictable factors, we focused on short- or medium-term (4- or 8-week) predictions. Second, as the data required to develop and evaluate the decision trees considered here are not available in the real world, we had to rely on simulated trajectories to synthetize the datasets needed to train and evaluate our decision trees. As discussed before, the factors driving the COVID-19 pandemic (e.g., public health responses, population behavior and adherence to mitigating strategies, seasonal effects on the transmission of SARS-CoV-2, and vaccination coverage) have continuously changed since the beginning of the pandemic and they will most likely continue to change. Hence, predictive models trained on historical data may not perform well when employed during the upcoming winter and spring. To mitigate this issue, we used simulated trajectories, which were selected to properly match the historical data and then projected over future months, to produce the datasets needed to train and evaluate our decision trees. This allowed us to account for a wide range of factors, and the uncertainties around them, that will derive the local trajectories of COVID-19 over the medium-term future (Table 2). While we estimated the accuracy, sensitivity, and specificity of each decision model using trajectories not included to train our decision trees, the actual performance of the proposed trees might be different when used in practice. The local policymakers who decide to use the decision trees proposed here are in the ideal position to measure the true accuracy, sensitivity, and specificity of these models using real-world data. Since such data is not currently available, the simulation approach we described here appear to be the only approach at the present to develop and evaluate the proposed decision rules.

Third, our simulation model did not differentiate vaccinated individuals based on the type of vaccine or the number of vaccine doses they have received. However, as none of the proposed decision tree models identified vaccine coverage as an important feature, relaxing this assumption is not expected to change our conclusions. Finally, in addition to surveillance systems we considered in our analysis (Table 1), data from other surveillance systems may also be available and used to provide information about different aspects of the pandemic. This may include genomic surveillance at hospitals to estimate the proportion of hospitalizations that are due to novel variants or potential vaccine-escape SARS-CoV-2 variants (23, 25), and seroprevalence surveillance to estimate the percentage of populations who have antibodies against SARS-CoV-2 (26). While including data from these sources could improve the performance of decision rules developed here, these sources are not always avilable at granular geographic regions.

Since the beginning of the COVID-19 pandemic, numerous models have been developed to predict the future trajectory of the pandemic (e.g., COVID-19 Forecast Hub (3) or the IHME COVID-19 Forecasting Model (4)). The results of these predictive models are usually available at the national or state levels. Therefore, the usefulness of these models for local policymakers are limited since the local trajectory of the pandemic could be substantially different from the predictions made at the larger geographic regions. The simple, easy-to-communicate decision rules we characterized in this study could be used to alert local policymakers when the hospital occupancy due to COVID-19 is to exceed the local hospital capacity.

While we validated these decision rules using trajectories under various scenarios, the true performance of these decision rules is to be seen. If the true accuracy, sensitivity, and specificity of the proposed decision rules turn out to be similar to what we estimated using simulated trajectories, the work presented here offers a novel and innovative approach to assist local policymakers in responding to future pandemics when real-word data to inform predictive and simulation models are scarce or not yet available.

## Data Availability

All data produced in the present work are contained in the manuscript.

## Acknowledgments

This work was partially funded by a Yale School of Public Health COVID-19 Rapid Response Grant to RY. RY was supported by R01AI153351.YHG was supported in part by contract 200-2016-91779 with the Centers for Disease Control and Prevention. JAS was supported by funding from the Centers for Disease Control and Prevention though the Council of State and Territorial Epidemiologists (NU38OT000297). The funders had no role in study design, data collection and analysis, decision to publish, or preparation of the manuscript.

## Disclaimer

The findings and conclusions in this report are those of the authors and do not necessarily represent the views of the National Institute of Allergy and Infectious Diseases or the Centers for Disease Control and Prevention

