## Supplement for "Simple decision rules to predict local surges in COVID-19 hospitalizations during the winter and spring of 2022"

### Supplementary Material: Additional Details on Simulation and Predictive Models

#### S1 Initialization of the simulation model

The model is initialized with a random number of susceptible individuals, drawn from the uniform distribution [250,000, 1,250,00]. The susceptible individuals are then randomly assigned according to the age groups 0-4, 5-12, 13-17, 18-29, 30-49, 50-64, 65-74, and 75+ according to the U.S. age-distribution (respectively, 6.0%, 10.0%, 6.4%, 16.3%, 25.7%, 19.2%, 9.6%, and 6.9%) (1). A number of infectious individuals, determined based on a random draw from the discrete-uniform (1, 5), are imported to Infectious compartments according to a Multinominal distribution with probabilities equal to the U.S. age distribution.

#### S2 Transmission model

##### S2.1 Simulation framework

To construct the model, we introduce the following notation:

- $i \in \{1, 2, 3, 4, 5, 6, 7, 8\}$ index of age groups {0-4, 5-12, 13-17, 18-29, 30-49, 50-64, 65-74, 75+}.
- $k \in \{1,2,3\}$ index of infection profile {infected with the ancestral strain, infected with the delta variant, infected with the novel variant}.
- $v \in \{0,1\}$ index of vaccination status
- $t$: epidemic time (in year).
- $N_i(t)$: number of individuals in age group $i$ at time $t$.
- $S_i(t)$: number of susceptible individuals in age group $i$ at time $t$.
- $V_i(t)$: number of vaccinated individuals in age group $i$ at time $t$.
- $E_{i,k,v}(t)$: number of exposed individuals in age group $i$ and with infection profile $k$ and vaccination status $v$ at time $t$.
- $I_{i,k,v}(t)$: number of infectious individuals in age group $i$ and with infection profile $k$ and vaccination status $v$ at time $t$.
- $H_{i,k,v}(t)$: number of hospitalized individuals in age group $i$ and with infection profile $k$ and vaccination status $v$ at time $t$.
- $R_{i,k,v}(t)$: number of recovered individuals in age group $i$ and with infection profile $k$ and vaccination status $v$ at time $t$.

The state of the epidemic at any given time $t$ can be identified by a discrete-time Markov chain $\{(S_i(t), V_i(t), E_{i,k,v}(t), I_{i,k,v}(t), H_{i,k,v}(t), R_{i,k,v}(t), i \in \{1,2, \dots ,8\}, k \in \{1, 2, 3\}, v \in \{0,1\}: t = 0, \Delta t, 2\Delta t, 3\Delta t, \dots\}$, where $\Delta t$ is the time-step of the simulation (e.g., $\Delta t = 1$ day). To generate epidemic trajectories for this model, we use Monte Carlo simulation to sample from this Markov chain using the following approach. Consider a particular compartment Z in which members depart due to $J$ events each of which is occurring at the rate $\mu_j, j \in \{1,2,\dots,J\}$ . For example, members of Susceptible compartment may leave due to 1) infection with the ancestral strain, 2) infection with the delta variant, 3) infection with the novel variant, or 4) vaccination (i.e., $J = 4$) (see Fig. 1). If the number of individuals in compartment Z at time $t$ is $Z(t)$, then the number of individuals that leave this compartment due to events $j \in \{1,2,\dots,J\}$ follows a multinomial distribution with total counts of $Z(t)$ and probabilities $(p_0, p_1, p_2, \dots p_J)$, where $p_0 = e^{-\sum_{j=1}^{J} \mu_j \Delta t}$ is the probability of not leaving the compartment Z during $[t, t + \Delta t]$, and $p_j = \frac{\mu_j}{\sum_{j=1}^{J} \mu_j \Delta t}(1 - p_0)$ is the probability of leaving the compartment Z during $[t, t + \Delta t]$ due to the event $j \in \{1,2,\dots,J\}$.

To identify the new epidemic state at the next time step, we first sample from the multinomial distributions associated to each compartment and then use these realizations to calculate the new epidemic state given the current epidemic state. The events that drive the epidemic are represented by black arrows in Fig. 1. For example, the number of susceptibles in age group $i$ at time $t + \Delta t$ can be calculated as:

$$\begin{aligned}
&S_i(t + \Delta t) = S_i(t) \\
&- \text{new infections with ancestral strain in age group } i \\
&- \text{new infections with the delta variant in age group } i \\
&- \text{new infections with novel variant in age group } i \\
&- \text{new vaccinations in age group } i \\
&+ \text{members losing infection-induced immunity in age group } i \\
&+ \text{members losing vaccine-induced immunity in age group } i.
\end{aligned}$$

#### S2.2 Rate of infection

For susceptible members in age group $i$, we calculate the rate of infection with ancestral strain at time $t$ as:

$$\mathcal{F}_{i,0}(t) = \beta(t) \sum_j \lambda_{i,j} \left( \frac{I_{j,0,0}(t)}{N_j(t)} + \frac{I_{j,0,1}(t)}{N_j(t)} \eta_{0,1} \right), \quad (1)$$

where $\beta(t)$ is the transmission parameter for the ancestral strain at time $t$, $\lambda_{i,j}$ is the daily rate at which an average individual in age group $i$ contact with individuals in age group $j$ (see below for how $\lambda_{i,j}$ is estimated), and $\eta_{0,1} \in [0,1]$ is the ratio of infectiousness for a vaccinated individual to an unvaccinated one who are infected with the ancestral strain.

For susceptible members in age group $i$, we calculate the rate of infection with the delta variant and a novel variant at time $t$ as:

$$\mathcal{F}_{i,1}(t) = \beta(t) \sum_j \lambda_{i,j} \eta_1 \left( \frac{I_{j,1,0}(t)}{N_j(t)} + \frac{I_{j,1,1}(t)}{N_j(t)} \eta_{1,1} \right), \quad (2)$$

$$\mathcal{F}_{i,2}(t) = \beta(t) \sum_j \lambda_{i,j} \eta_2 \left( \frac{I_{j,2,0}(t)}{N_j(t)} + \frac{I_{j,2,1}(t)}{N_j(t)} \eta_{2,1} \right), \quad (3)$$

Where $\eta_1 \geq 0$ and $\eta_2 \geq 0$ are the ratio of infectiousness for the delta and the novel variant to the ancestral strain, and $\eta_{1,1} \in [0,1]$ and $\eta_{2,1} \in [0,1]$ are the ratio of infectiousness for a vaccinated individual who is infected with the delta variant and the novel variant to the unvaccinated individual who is infected with the ancestral strain.

For vaccinated members in age group $i$, we calculate the rate of infection with variant $k \in \{1,2,3\}$ at time $t$ as:

$$\mathcal{F}_{i,k}^{V}(t) = (1-\gamma_k)\mathcal{F}_{i,k}(t), \tag{4}$$

where $\gamma_k \in [0,1]$ is the effectiveness of vaccination in providing immunity against infection with the variant $k$.

To capture the effect of seasonal changes on the transmission of SARS-CoV-2, we allow the transmission parameter $\beta(t)$ to vary over time according to:

$$\beta(t) = a_0 + a_1 \cos 2\pi(t-\phi)\,. \tag{5}$$

Here, $a_0$ is the baseline transmissibility which is not influenced by the seasonality effect, and the parameter $a_1$ represents the maximum magnitude of seasonality effect during a year. The phase parameter $\phi$ determines when the effect of seasonality reaches its maximum of minimum and is included to provide additional flexibility in the modeling of the seasonality effect. We determined that $a_0$, $a_1$, and $\phi$ by random draws from uniform distributions $U[0.75, 1.25]$, $U[0, 0.5]$, and $U[-0.25, 0]$. See Fig. S4 for how $\beta(t)$ behaves for different values of $a_0$, $a_1$, and $\phi$.

#### S2.3 Daily contact rate

Let $\hat{\lambda}_{i',j'}$ denote the average number of contacts per day an individual in age group $j'$ makes with individuals in age group $i'$, as estimated by Prem et al. (2) for the U.S. population. Age groups considered in this survey include $\hat{\mathcal{A}}$ = {0-4, 5-9, 10-14, 5-19, 20-24, 25-29, 30-34, 35-39, 40-44, 45-49, 50-54, 55-59, 60-64, 65-69, 70, 75+}.

Since our model only includes age groups $\mathcal{A}$ = {0-4, 5-12, 13-17, 18-29, 30-49, 50-64, 65-74, 75+}, we use estimates $\hat{\lambda}_{i',j'}$, $i', j' \in \hat{\mathcal{A}}$, to calculate the contact matrix $[\lambda_{i,j}]$, $i, j \in \mathcal{A}$ to be used in our model. To this end, we make the following assumption:

1. Contacts *within* an age group are uniformly distributed; that is, a contact with an average member in age group {0-4}, for example, is equally likely to be with someone of age 0, 1, 2, 3, or 4.
2. If in estimating $\lambda_{i,j}$, the age group $j \in \mathcal{A}$ includes multiple age groups from $\hat{\mathcal{A}}$ (say $j'_1, j'_2$, ...), then $\lambda_{i,j} = \sum_{j' \in \{j'_1, j'_2, \ldots\}} \hat{\lambda}_{i',j'}$. For example, $\lambda_{\{35-39\},\{64-74\}} = \hat{\lambda}_{\{35-39\},\{64-69\}} + \hat{\lambda}_{\{35-39\},\{70-74\}}$.
3. If in estimating $\lambda_{i,j}$, age group $i \in \mathcal{A}$ overlaps with multiple age groups from $\hat{\mathcal{A}}$ (say $i'_1, i'_2$, ...), the daily contact $\lambda_{i,j}$ will be the average of $\lambda_{i'_1,j}$, $\lambda_{i'_2,j}$, .... For example, $\lambda_{\{30-49\},\{0-4\}} = (\hat{\lambda}_{\{30-34\},\{0-4\}} + \hat{\lambda}_{\{35-39\},\{0-4\}} + \hat{\lambda}_{\{40-44\},\{0-4\}} + \hat{\lambda}_{\{45-49\},\{0-4\}})/4$.

Following the approach of Medlock et al. (3), we then ensured that the number of contacts between age groups is symmetric (i.e. $\varpi_i \lambda_{i,j} = \varpi_j \lambda_{j,i}$, where $\varpi_i$ is the size of age group $i$), by using $\bar{\lambda}_{i,j} = \frac{1}{2\varpi_i}(\varpi_i \lambda_{i,j} + \varpi_j \lambda_{j,i})$. The contact rate matrix $[\lambda_{i,j}]$ used in our model is shown in Table S1.

Table S1: Average daily number of contacts between different age groups used in our model

| Age Groups | 0-4 | 5-12 | 13-17 | 18-29 | 30-49 | 50-64 | 65-74 | 75+ |
|---|---|---|---|---|---|---|---|---|
| **0-4** | 2.598 | 1.312 | 0.316 | 1.146 | 2.551 | 1.080 | 0.258 | 0.141 |
| **5-12** | 0.786 | 6.398 | 2.266 | 1.069 | 3.280 | 1.306 | 0.365 | 0.207 |
| **13-17** | 0.295 | 3.533 | 6.294 | 5.178 | 3.624 | 1.687 | 0.296 | 0.216 |
| **18-29** | 0.419 | 0.652 | 2.026 | 6.492 | 4.244 | 1.944 | 0.230 | 0.104 |
| **30-49** | 0.591 | 1.268 | 0.899 | 2.691 | 6.790 | 2.366 | 0.412 | 0.195 |
| **50-64** | 0.336 | 0.678 | 0.562 | 1.655 | 3.176 | 2.944 | 0.476 | 0.173 |
| **65-74** | 0.161 | 0.379 | 0.197 | 0.392 | 1.105 | 0.951 | 1.144 | 0.253 |
| **75+** | 0.122 | 0.299 | 0.200 | 0.247 | 0.729 | 0.482 | 0.352 | 0.396 |

##### S2.4 Effectiveness of control measures

We assume that control measures went into effect whenever the rate of hospital occupancy due to COVID-19 exceeded the threshold $T_1$ and were lifted whenever this rate dropped below the second threshold $T_2$ (4). These thresholds differ across simulated trajectories and were determined by random draws from uniform distributions $U(1, 50)$ and $U(1, 50)$ per 100,000 population. We assume that the impact of control measures ($\alpha(h)$) in reducing the effective reproductive number varies as a function of hospital occupancy due to COVID-19 ($h$) according to the sigmoid function $\alpha(h) = \frac{\bar{\alpha}}{1+e^{-4h/\bar{h}}}$, where $\bar{\alpha}$ is the maximum impact of control measures and $\bar{h}$ is the maximum hospitalization capacity that could be allocated for COVID-19 patients (Fig. S3). We determined $\bar{\alpha}$ and $\bar{h}$ by random draws from uniform distributions $U(50\%, 85\%)$ (4-6) and $U(5, 15)$ per 100,000 population (7).

#### S3 Selection of simulated trajectories

To ensure that the trajectories simulated by our model are consistent with the spread of SARS-CoV-2 in the U.S., we only considered trajectories that satisfied certain conditions related to the following outcomes.

1. **Weekly rate of hospital occupancy associated with COVID-19**: We only considered trajectories where the weekly hospital occupancy rate reaches at least 1.1 per 100,000 population but does not surpass 61.1 per 100,000 population. These thresholds are informed by hospital occupancy associated with COVID-19 in U.S. states during the period April 1 and July 7, 2020 (8)
2. **Weekly hospitalizations rates**: We only considered trajectories where the weekly rate of new hospitalizations reaches at least $T_1$ but does not surpass $T_2$ per 100,000 population, where $T_1$ is 0.75 times the minimum rate of new hospitalizations and $T_2$ is 1.25 times the maximum rate of new hospitalizations observed in the surveillance sites of COVID-Net (Table S7).
3. **Prevalence of population with immunity from infection**: We only considered trajectories where the prevalence of population with immunity from infection does not surpass 35%, as informed by the CDC's seroprevalecnce survey (9). This seroprevalecnce survey estiamtes that on average 20.6% of the U.S. population had immunity from infection on Auguest 26, 2021 with the state-level mimimum of 1.6% and the maximum of 34.1%. To measure how well a simulated trajectory matches these estimates, we estimate

the likelihood of observing the seroprevalence of $\hat{\mu} = 20.6\%$, if the simulation trajectory results in the seroprevalence of $\mu$ using:

$$L_1 = f(x = \mu; \hat{\mu}, \hat{\sigma}),$$

where $f$ is the probability density function of a normal distribution with mean $\hat{\mu}$ and standard deviation of $\hat{\sigma} = (34.1\% - 1.6\%)/4$.

4. **Cumulative hospitalization rate**: To measure how well a simulated trajectory matches the observed data on cumulative hospitalization rate (i.e., the overall cumulative hospitalization rate of 768.0 per 100,000 population, with minimum of 301.7 and maximum of 1050.3 observed in the states included in COVID-NET, Table S7), we calculate the livelihood of this observation assuming that the simulated trajectory represents the reality. To this end, we measure the likelihood of observing the cumulative hospitalization rate of $\hat{\mu} = 768.0$ per 100,000 population, if the simulation trajectory results in the cumulative hospitalization rate of $\mu$ using:

$$L_2 = f(x = \mu; \hat{\mu}, \hat{\sigma}),$$

where $f$ is the probability density function of a normal distribution with mean $\hat{\mu}$ and standard deviation of $\hat{\sigma} = (1050.3\ - 301.7)/4$.
5. **Cumulative hospitalization rate by age**: We used the same approach as described above to calculate the likelihood of observing hospitalization rates in each age group, as reported in Table S7. This returns likelihoods $L_{3,1}, L_{3,2}, \ldots, L_{3,8}$ for 8 age groups included in our model.
6. **Cumulative vaccination rate**: We used the same approach as described above to calculate the likelihood ($L_4$) of observation related to vaccination rates as reported in Table S9.
7. **Prevalence of the delta variant among new infections**: We used the same approach describe above to calculate the likelihood ($L_5$) of observations related to the prevalence of the delta variant among new infections (Table S10).

We calculate the natural logarithm of the likelihood of a trajectory as:

$$\ln \mathcal{L} = \ln L_1 + \ln L_2 + \sum_{k=1}^{8} \ln L_{3,k} + \ln L_4 + \ln L_5.$$

To build a set of trajectories to train predictive models, we simulated as many trajectories as needed to obtain 7,500 feasible trajectories. For each simulated trajectory, parameter values are randomly drawn from the probability distribution of epidemic parameters listed in Table S2-Table S5. These prior distributions are informed by estimates extracted from existing scientific literature when such estimates are available; when not available, we assumed biologically-feasible distributions. Among the total of 293,193 simulated trajectory, we discarded 285,693 trajectories that violated the feasibility conditions described above and calculated the above pseudo-likelihood function for the remaining trajectories. After calculating $\ln \mathcal{L}$ for each simulation trajectory, we used 2,000 trajectories randomly selected among trajectories with a positive $\ln \mathcal{L}$ to train the predictive models.

### S4 Training decision tree models

We used the scikit-learn package to train decision tree models (10). Before training the models, we created a balanced dataset where the number of observations for which the hospital occupancy passed the specified threshold is equal to the number of observations for which this event didn't occur. To avoid overfitting, we used a minimal cost-complexity pruning approach (11), where we determined the complexity parameter using 10-fold cross-validation to maximize the model accuracy (defined as the fraction of correct predictions) (12). For each decision tree model, we chose the optimal value of the complexity parameter from {0, 0.005, 0.01, 0.015, 0.02, …, 0.1}.

### S5 Supplementary Tables

Table S2: Model parameters related to dominant strain

| Parameter | Prior Distribution[#] | 95% Percentile Interval[†] | Source |
|---|---|---|---|
| $R_0$ | Beta (mean=2.5, SD=0.75, min=1.5, max=4) | (1.52, 3.85) | (13, 14) |
| Delay from infection until becoming infectious (days) | Beta (mean=5, SD=0.5, min=4, max=6) | (4.06, 5.96) | (15) |
| Duration of infectiousness (days) | Beta (mean=4, SD=0.5, min=2, max=8) | (2.05, 7.22) | (16, 17) |
| Duration of hospitalization (days) | Beta (mean=12, SD=1, min=7, max=17) | (10.06, 13.94) | (18) |
| Duration of infection-induced immunity (years) | Beta (mean=1, SD=0.25, min=0.25, max=1.5) | (0.493, 1.42) | (6, 19) |
| Relative probability of hospitalization for the dominant strain | | | ⁋ |
| 0-4 | Gamma (mean=0.5, SD=0.1) | | |
| 5-12 | Gamma (mean=0.5, SD=0.1) | | |
| 13-17 | Gamma (mean=0.25, SD=0.05) | | |
| 18-29 | Reference group | | |
| 30-49 | Gamma (mean=2.0, SD=0.5) | | |
| 50-64 | Gamma (mean=4.0, SD=1.0) | | |
| 65-74 | Gamma (mean=6.0, SD=1.5) | | |
| 75+ | Gamma (mean=10.8, SD=2.7) | | |
| Probability of hospitalization for age group 18-29 if infected | Uniform (0.0005, 0.005) | | Assumption |
| Probability death if requiring hospitalization (%) | | | (20) |
| 0-4 | Beta (mean=0.2, SD=0.05) | | |
| 5-12 | Beta (mean=0.2, SD=0.05) | | |
| 13-17 | Beta (mean=0.2, SD=0.05) | | |
| 18-29 | Beta (mean=2.6, SD=0.65) | | |
| 30-49 | Beta (mean=2.6, SD=0.65) | | |
| 50-64 | Beta (mean=7.9, SD=1.98) | | |
| 65-74 | Beta (mean=14.1, SD=3.53) | | |
| 75+ | Beta (mean=20.9, SD=5.23) | | |
| Weekly rate of importation of new infection | 5 | | Assumption |

[#] SD: Standard deviation

[†] 95% Percentile Interval is defined as [2.5th percentile, 97.5th percentile] of the assumed probability distribution.

⁋ Risk for COVID-19 Infection, Hospitalization, and Death by Age Group (https://www.cdc.gov/coronavirus/2019-ncov/covid-data/investigations-discovery/hospitalization-death-by-age.html).

Table S3: Model parameters related to novel variant

| Parameter | Distribution# | Source |
|---|---|---|
| Weekly importation rate of the delta variant ($\gamma(t) = b_{max}/(1 + e^{-b(t-t_{mid})}$), see Fig. S1) | | Assumption |
| $b$ | Beta (mean=7, SD=0.5, min=5, max=9) | |
| $b_{max}$ | Uniform (2, 5) | |
| $t_{mid}$ | Uniform (1.5, 1.75) | |
| Importation rate of the novel variant ($\gamma(t) = b_{max}/(1 + e^{-b(t-t_{mid})}$), see Fig. S1) | | Assumption |
| $b$ | Beta (mean=7, SD=0.5, min=5, max=9) | |
| $b_{max}$ | Uniform (0, 5) | |
| $t_{mid}$ | Uniform (1.5, 2) | |
| Delay from infection until becoming infectious (days) | Same distribution as dominant stain in Table S2 | Assumption |
| Duration of hospitalization (days) | Same distribution as dominant stain in Table S2 | Assumption |
| Duration of infection-induced immunity (years) | Same distribution as dominant stain in Table S2 | Assumption |
| Ratio of transmissibility to that of the dominant strain | Uniform (1, 2) | (21, 22) |
| Ratio of the duration of infectiousness for the novel strain to that of the dominant strain | Uniform (0.5, 2) | Assumption |
| Ratio of the probability of requiring hospitalization to that of the dominant strain | Uniform (0.5, 3) | (23-25) |
| Probability death if requiring hospitalization (%) | The same as dominant stain in Table S2. | Assumption |
| Effectiveness of the recent recovery from infection with the dominant strain in providing immunity against infection with the novel strain | Uniform (50%, 100%) | Assumption |

Table S4: Model parameters related to vaccinated individual who are infected with the novel variant

| Parameter | Distribution[#] | Source |
|---|---|---|
| Delay from infection until becoming infectious (days) | Same distribution as dominant stain in Table S2 | Assumption |
| Duration of hospitalization (days) | Same distribution as dominant stain in Table S2 | Assumption |
| Duration of infection-induced immunity (years) | Same distribution as dominant stain in Table S2 | Assumption |
| Vaccine effectiveness against novel variant | | (26) |
| Ratio of susceptibility of a vaccinated individual that of an unvaccinated individual | Uniform (0, 1) | |
| Ratio of transmissibility to that of the dominant strain | Uniform (0, 0.5) | |
| Ratio of duration of infectiousness for the vaccinated individuals to that of the dominant strain | Uniform (0, 0.5) | |
| Ratio of the probability of requiring hospitalization for the vaccinated individuals to that of the dominant strain | Uniform (0, 0.5) | |
| Probability death if requiring hospitalization (%) | The same as dominant stain in Table S2. | |

Table S5: Model parameters related to vaccine and vaccination

| Parameter | Distribution[#] | Source |
|---|---|---|
| Duration of vaccine-induced immunity, accounting for booster doses (years) | Uniform (0.5, 2.5) | (6) |
| Increase in the duration of infection-induced immunity after vaccination | Uniform (0%, 50%) | Assumption |
| Vaccine effectiveness against infection | | Assumption based on (22) |
| For dominant strain | Uniform (0%, 100%) | |
| For novel strain | Uniform (0%, 100%) | |
| Vaccine effectiveness against hospitalization | | Assumption based on (22, 27) |
| For dominant strain | Uniform (85%, 100%) | |
| For novel strain | Uniform (0%, 100%) | |
| Vaccine effectiveness in reducing infectiousness | | Assumption based on (28) |
| For dominant strain | Uniform (25%, 75%) | |
| For novel strain | Uniform (25%, 75%) | |
| Annual vaccination rate $\boldsymbol{v_k(t)} = b_{max} + (b_{max} - b_{min})/(1 + e^{-b(t-t_{mid}-t_{min})})$ if $t \geq t_{min}$ (see Fig. S2) | | Assumption to match the estimated vaccination coverage (Table S8) |
| $b$ | Uniform (-20, -10) | |
| $b_{min}$ | 0 | |
| $t_{mid}$ | Uniform (0.25, 0.75) | |
| $b_{max}$ | Uniform (2, 3) | |
| $b_{max}$ | Uniform (0.25, 0.75) | |
| 5-12 | Uniform (1, 3) | |
| 13-17 | Uniform (1, 3) | |
| 18-29 | Uniform (1, 3) | |
| 30-49 | Uniform (1, 3) | |
| 50-64 | Uniform (2, 4) | |
| 65-74 | Uniform (3, 5) | |
| 75+ | Uniform (2, 4) | |
| $t_{min}$ | | |
| 5-12 | Uniform (1.6, 1.8) | |
| 13-17 | Uniform (1.0, 1.4) | |
| 18-29 | Uniform (0.85, 1.25) | |
| 30-49 | Uniform (0.85, 1.25) | |
| 50-64 | Uniform (0.85, 1.25) | |
| 65-74 | Uniform (0.75, 1.15) | |
| 75+ | Uniform (0.7, 1.1) | |

[#] SD: Standard deviation

[†] 95% Percentile Interval is defined as [2.5th percentile, 97.5th percentile] of the assumed probability distribution.

Table S6: Model parameters related to mitigating strategies

| Parameter | Distribution[#] | Source |
|---|---|---|
| Effectiveness of control measures ($\alpha(h) = \frac{\bar{\alpha}}{1+e^{-4h/\bar{h}}}$) | | |
| Maximum effectiveness of control measures ($\bar{\alpha}$) | $U(50\%, 85\%)$ | (4-6) |
| Maximum hospitalization capacity that could be allocated to patients with COVDI-19, per 100,000 population ($\bar{h}$) | $U(5, 15)$ | (7) |

Table S7: Summary of COVID-NET hospitalization data to inform selection of simulated trajectories.

| **Weekly hospitalization rate (per 100,000 population) between March 1, 2010 and November 27, 2021** (source: https://gis.cdc.gov/grasp/COVIDNet/COVID19_3.html) | | | |
|---|---|---|---|
| | | Minimum value achieved in all States included in COVID-NET | Maximum value achieved in all States included in COVID-NET |
| Overall | | 8.9 | 51.2 |
| **Cumulative hospitalization rate (per 100,000 population) between March 1, 2010 and November 27, 2021** (source: https://gis.cdc.gov/grasp/COVIDNet/COVID19_3.html) | | | |
| | Cumulative Hospitalization Rate | Minimum value among all States included in COVID-NET | Maximum value among all States included in COVID-NET |
| Overall | 768.0 | 301.7 | 1050.3 |
| 0-4 | 100.2 | 50.6 | 161.2 |
| 5-11 | 33.7 | 17.4 | 46.6 |
| 12-17 | 81.4 | 43.5 | 129.7 |
| 18-29 | 298.8 | 85.0 | 508.0 |
| 30-49 | 601.5 | 179.3 | 897.0 |
| 50-64 | 1124.2 | 449.0 | 1490.7 |
| 65-74 | 1590.9 | 750.1 | 2356.8 |
| 75+ | 2973.2 | 1553.9 | 4313.6 |

Table S8: Estimated vaccine coverage (i.e., the percentage of the population that are fully vaccinated) over time by age (source: https://covid.cdc.gov/covid-data-tracker/#vaccination-demographics-trends)

| **Date** | **Overall** | **0-4** | **5-12** | **13-17** | **18-29** | **30-49** | **50-64** | **65-75** | **75+** |
|---|---|---|---|---|---|---|---|---|---|
| Jan 1, 2021 | 0.01% | 0.00% | 0.00% | 0.00% | 0.01% | 0.02% | 0.02% | 0.02% | 0.01% |
| Jan 26, 2021 (week 4) | 1.72% | 0.00% | 0.00% | 0.02% | 1.53% | 2.63% | 2.56% | 1.68% | 1.94% |
| Feb 26, 2021 (week 8) | 9.39% | 0.00% | 0.01% | 0.13% | 5.39% | 8.51% | 9.88% | 21.22% | 32.80% |
| Mar 26, 2021 (week 12) | 18.79% | 0.00% | 0.01% | 0.36% | 9.34% | 14.58% | 20.31% | 53.66% | 60.95% |
| Apr 23, 2021 (week 16) | 33.75% | 0.00% | 0.01% | 2.09% | 21.65% | 30.79% | 47.94% | 76.76% | 75.76% |
| May 21, 2021 (week 20) | 45.70% | 0.00% | 0.02% | 9.14% | 38.53% | 47.83% | 64.78% | 83.93% | 80.63% |
| June 18, 2021 (week 24) | 51.65% | 0.00% | 0.34% | 26.34% | 46.26% | 54.87% | 70.60% | 86.92% | 82.75% |
| July 16, 2021 (week 28) | 54.31% | 0.00% | 0.44% | 34.16% | 49.93% | 57.99% | 73.02% | 88.27% | 83.70% |
| Aug 13, 2021 (week 32) | 56.46% | 0.00% | 0.45% | 39.98% | 52.94% | 60.55% | 75.09% | 89.45% | 84.51% |
| Sep 10, 2021 (week 36) | 59.74% | 0.00% | 0.45% | 48.54% | 57.25% | 64.70% | 78.22% | 91.22% | 85.72% |
| Oct 8, 2021 (week 40) | 62.44% | 0.00% | 0.45% | 53.62% | 61.25% | 68.41% | 80.69% | 92.83% | 86.91% |
| Nov 5, 2021 (week 44) | 64.13% | 0.00% | 0.46% | 56.01% | 63.61% | 70.66% | 82.29% | 94.32% | 88.24% |
| Dec 3, 2021 (week 48) | 65.74% | 0.00% | 4.63% | 57.52% | 65.21% | 72.26% | 83.57% | 95.42% | 89.24% |

We note that the age groups in our model is different from the age groups of COVID Data Tracker. To estimate the vaccine coverage for age-group 5-12, we divide the total number of vaccinations by 2/3 the population size of in age groups <12. To estimate the vaccine coverage for age group 13-17, we divide the number of vaccinations in age group 12-15 and 16-17 by the population sizes of the age groups. To estimate the vaccine coverage for age group 18-29, we divided the total number of vaccinations in age group 18-24 and 1/3 in age group 25-39 by the same proportions of the population sizes of the age groups. To estimate the vaccine coverage for age group 30-49, we divided 2/3 of vaccinations in age group 25-39 and the total number in age group 40-49 by the same proportions of the population sizes of the age groups.

Table S9: Estimated vaccination coverage (i.e., the percentage of the population that are fully vaccinated) (source: https://github.com/govex/COVID-19/tree/master/data_tables/vaccine_data)

| **Rate of vaccination on December 07, 2021** | | | |
|---|---|---|---|
| | Mean | Minimum value achieved in all States | Maximum value achieved in all State |
| Overall | 59.83% | 46.21% | 80.64% |

Table S10: Percentage of cases due to delta variant https://covid.cdc.gov/covid-data-tracker/#variant-proportions

| Last Day of 2-week Sequencing | USA | Minimum value observed among 10 U.S. regions | Maximum value observed among 10 U.S. regions |
|---|---|---|---|
| 4/10/2021 | 0.1% | 0.0% | 0.3% |
| 4/24/2021 | 0.6% | 0.3% | 1.2% |
| 5/8/2021 | 1.4% | 0.8% | 2.4% |
| 5/15/2021 | 2.4% | 1.2% | 7.5% |
| 5/22/2021 | 3.8% | 2.1% | 13.9% |
| 5/29/2021 | 7.0% | 2.3% | 23.6% |
| 6/5/2021 | 14.1% | 5.1% | 44.7% |
| 6/12/2021 | 26.1% | 4.9% | 70.6% |
| 6/19/2021 | 37.2% | 16.3% | 77.7% |
| 6/26/2021 | 53.6% | 28.1% | 87.8% |
| 7/3/2021 | 69.4% | 51.9% | 93.0% |
| 7/10/2021 | 82.2% | 70.9% | 95.9% |
| 7/17/2021 | 87.6% | 82.6% | 95.9% |
| 7/24/2021 | 94.4% | 93.1% | 97.0% |
| 7/31/2021 | 96.8% | 93.8% | 99.0% |
| 8/7/2021 | 97.9% | 96.4% | 99.6% |
| 8/14/2021 | 98.8% | 97.7% | 99.8% |

Table S11: Correlations between the features defined in Table 1 and the event that hospital capacity would surpass the specified thresholds within 8 weeks.

| | | **Threshold for Hospital Occupancy (per 100,000 population)** | | | | | |
|---|---|---|---|---|---|---|---|
| | | **10** | | **15** | | **20** | |
| **Surveillance** | **Features used for prediction** | **Corr** | ***p*-value** | **Corr** | ***p*-value** | **Corr** | ***p*-value** |
| Rate of hospital occupancy | Current value | 0.507 | <0.001 | 0.613 | <0.001 | 0.678 | <0.001 |
| Weekly rate of new hospitalizations | Average over the past 2 weeks | 0.464 | <0.001 | 0.549 | <0.001 | 0.595 | <0.001 |
| | Average change during the past 4 weeks | 0.074 | <0.001 | 0.114 | <0.001 | 0.151 | <0.001 |
| Vaccination coverage | Cumulative value | -0.083 | <0.001 | -0.11 | <0.001 | -0.118 | <0.001 |
| Prevalence of novel variant among new infections | Average over the past 2 weeks | 0.023 | 0.050 | 0.028 | 0.02 | 0.011 | 0.348 |
| | Average change during the past 4 weeks | 0.032 | 0.008 | 0.049 | <0.001 | 0.035 | 0.003 |

Table S12: Correlations between the features defined in Table 1 and the event that hospital capacity would surpass the specified thresholds within 4 weeks.

| | | **Threshold for Hospital Occupancy (per 100,000 population)** | | | | | |
|---|---|---|---|---|---|---|---|
| | | **10** | | **15** | | **20** | |
| **Surveillance** | **Features used for prediction** | **Corr** | ***p*-value** | **Corr** | ***p*-value** | **Corr** | ***p*-value** |
| Rate of hospital occupancy | Current value | 0.598 | <0.001 | 0.71 | <0.001 | 0.767 | <0.001 |
| Weekly rate of new hospitalizations | Average over the past 2 weeks | 0.552 | <0.001 | 0.639 | <0.001 | 0.67 | <0.001 |
| | Average change during the past 4 weeks | 0.112 | <0.001 | 0.167 | <0.001 | 0.226 | <0.001 |
| Vaccination coverage | Cumulative value | -0.071 | <0.001 | -0.094 | <0.001 | -0.098 | <0.001 |
| Prevalence of novel variant among new infections | Average over the past 2 weeks | 0.025 | 0.019 | 0.026 | 0.015 | 0.029 | 0.006 |
| | Average change during the past 4 weeks | 0.035 | 0.001 | 0.043 | <0.001 | 0.047 | <0.001 |

Table S13: Performance of Decision Trees A and B (Fig. S6; 8-week prediction for the hospital occupancy threshold of 10 per 100,000 population) evaluated using 500 simulated trajectories not used for training these models.

| | **Accuracy** | **Sensitivity** | **Specificity** |
|---|---|---|---|
| **Base scenario** | | | |
| Decision Tree A | 0.918 | 0.871 | 0.928 |
| Decision Tree B | 0.917 | 0.785 | 0.947 |
| **If non-pharmaceutical measures are removed during the winter and spring of 2022** | | | |
| Decision Tree A | 0.965 | 0.819 | 0.974 |
| Decision Tree B | 0.969 | 0.733 | 0.984 |
| **If no novel variant emerges during the winter and spring of 2022** | | | |
| Decision Tree A | 0.911 | 0.852 | 0.924 |
| Decision Tree B | 0.915 | 0.834 | 0.933 |
| **Genomic surveillance with small sample size (*N*=250 tests per week)** | | | |
| Decision Tree A | 0.918 | 0.871 | 0.928 |
| Decision Tree B | 0.917 | 0.782 | 0.947 |

Table S14: Performance of Decision Trees A and B (Fig. S7; 8-week prediction for the hospital occupancy threshold of 20 per 100,000 population) evaluated using 500 simulated trajectories not used for training these models.

| | **Accuracy** | **Sensitivity** | **Specificity** |
|---|---|---|---|
| **Base scenario** | | | |
| Decision Tree A | 0.858 | 0.943 | 0.794 |
| Decision Tree B | 0.863 | 0.903 | 0.833 |
| **If non-pharmaceutical measures are removed during the winter and spring of 2022** | | | |
| Decision Tree A | 0.941 | 0.875 | 0.954 |
| Decision Tree B | 0.943 | 0.822 | 0.967 |
| **If no novel variant emerges during the winter and spring of 2022** | | | |
| Decision Tree A | 0.874 | 0.919 | 0.83 |
| Decision Tree B | 0.874 | 0.886 | 0.863 |
| **Genomic surveillance with small sample size (*N*=250 tests per week)** | | | |
| Decision Tree A | 0.858 | 0.943 | 0.794 |
| Decision Tree B | 0.863 | 0.903 | 0.833 |

Table S15: Performance of Decision Trees A and B (Fig. S8; 4-week prediction for the hospital occupancy threshold of 10 per 100,000 population) evaluated using 500 simulated trajectories not used for training these models.

| | **Accuracy** | **Sensitivity** | **Specificity** |
|---|---|---|---|
| **Base scenario** | | | |
| Decision Tree A | 0.946 | 0.930 | 0.952 |
| Decision Tree B | 0.946 | 0.930 | 0.952 |
| **If non-pharmaceutical measures are removed during the winter and spring of 2022** | | | |
| Decision Tree A | 0.98 | 0.933 | 0.985 |
| Decision Tree B | 0.98 | 0.933 | 0.985 |
| **If no novel variant emerges during the winter and spring of 2022** | | | |
| Decision Tree A | 0.952 | 0.955 | 0.952 |
| Decision Tree B | 0.952 | 0.955 | 0.952 |
| **Genomic surveillance with small sample size (*N*=250 tests per week)** | | | |
| Decision Tree A | 0.946 | 0.930 | 0.952 |
| Decision Tree B | 0.946 | 0.930 | 0.952 |

Table S16: Performance of Decision Trees A and B (Fig. S9; 4-week prediction for the hospital occupancy threshold of 15 per 100,000 population) evaluated using 500 simulated trajectories not used for training these models.

| | **Accuracy** | **Sensitivity** | **Specificity** |
|---|---|---|---|
| **Base scenario** | | | |
| Decision Tree A | 0.934 | 0.93 | 0.937 |
| Decision Tree B | 0.934 | 0.93 | 0.937 |
| **If non-pharmaceutical measures are removed during the winter and spring of 2022** | | | |
| Decision Tree A | 0.976 | 0.931 | 0.986 |
| Decision Tree B | 0.976 | 0.931 | 0.986 |
| **If no novel variant emerges during the winter and spring of 2022** | | | |
| Decision Tree A | 0.941 | 0.93 | 0.949 |
| Decision Tree B | 0.941 | 0.93 | 0.949 |
| **Genomic surveillance with small sample size (*N*=250 tests per week)** | | | |
| Decision Tree A | 0.934 | 0.93 | 0.937 |
| Decision Tree B | 0.934 | 0.93 | 0.937 |

Table S17: Performance of Decision Trees A and B (Fig. S10; 4-week prediction for the hospital occupancy threshold of 20 per 100,000 population) evaluated using 500 simulated trajectories not used for training these models.

| | **Accuracy** | **Sensitivity** | **Specificity** |
|---|---|---|---|
| **Base scenario** | | | |
| Decision Tree A | 0.924 | 0.967 | 0.873 |
| Decision Tree B | 0.924 | 0.967 | 0.873 |
| **If non-pharmaceutical measures are removed during the winter and spring of 2022** | | | |
| Decision Tree A | 0.966 | 0.948 | 0.973 |
| Decision Tree B | 0.966 | 0.948 | 0.973 |
| **If no novel variant emerges during the winter and spring of 2022** | | | |
| Decision Tree A | 0.932 | 0.962 | 0.889 |
| Decision Tree B | 0.932 | 0.962 | 0.889 |
| **Genomic surveillance with small sample size (*N*=250 tests per week)** | | | |
| Decision Tree A | 0.924 | 0.967 | 0.873 |
| Decision Tree B | 0.924 | 0.967 | 0.873 |

#### S6 Supplementary Figures

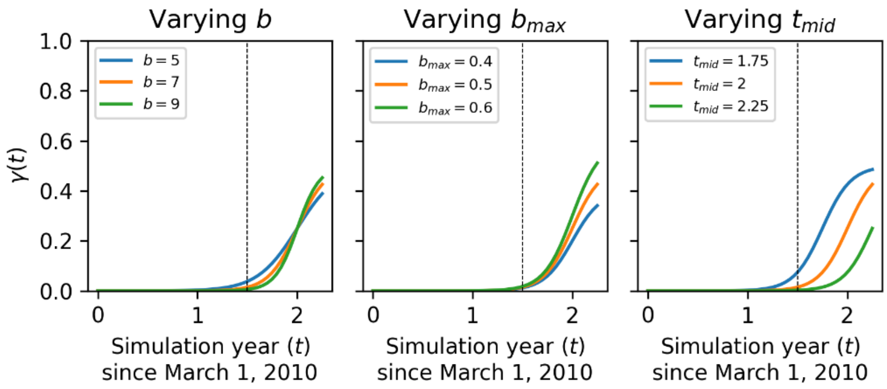


**Fig. S1: Probability that an imported case is infected with the novel strain.** We assumed that this probability increases according to $\boldsymbol{\gamma(t) = b_{max}/(1 + e^{-b(t-t_{mid})})}$. The probability distributions assumed for $\boldsymbol{b_{max}}$, $\boldsymbol{b}$, and $\boldsymbol{t_{mid}}$ are listed in Table S3. The vertical dotted lines marks September 1, 2021.

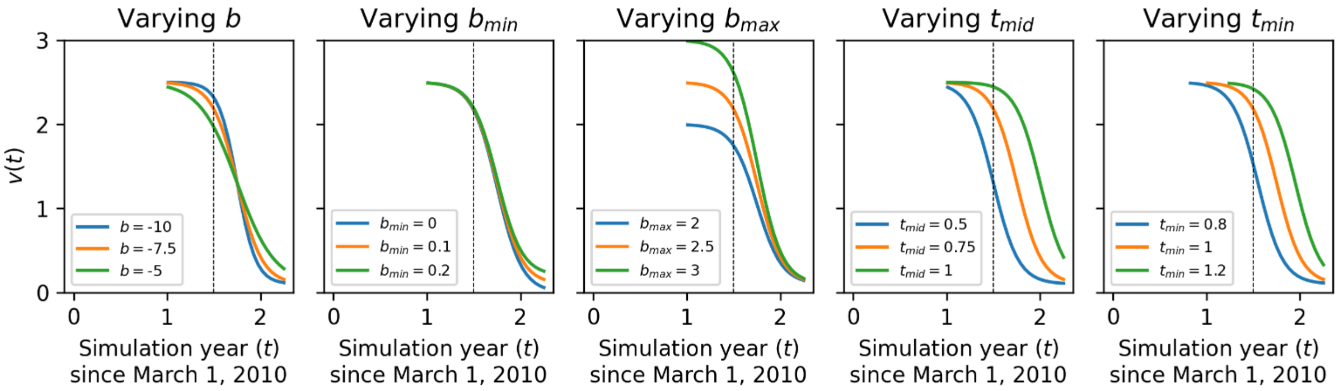


**Fig. S2: The rate at which population is getting vaccinated.** We assumed that vaccination rate in an age group decreases according to $\boldsymbol{v(t) = b_{max} + (b_{max} - b_{min})/(1 + e^{-b(t-t_{mid}-t_{min})})}$ if $\boldsymbol{t \geq t_{min}}$. The probability distributions assumed for $\boldsymbol{b_{max}}$, $\boldsymbol{b_{min}}$, $\boldsymbol{b}$, $\boldsymbol{t_{mid}}$, and $\boldsymbol{t_{min}}$ are listed in Table S5. The vertical dotted lines marks September 1, 2021.

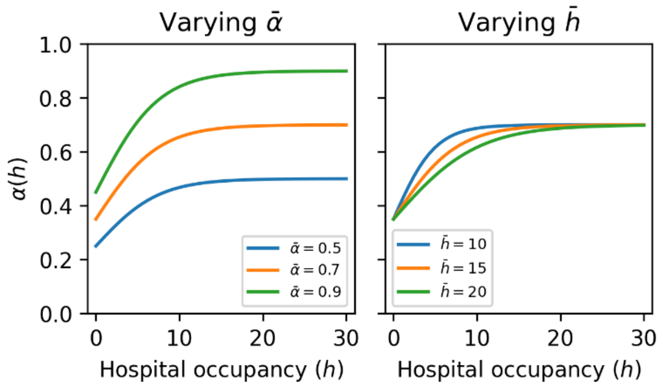


**Fig. S3: The effectiveness of control measures as the function of hospital occupancy.** We assumed that the effectiveness of control strategies increases with the hospital occupancy according to $\boldsymbol{\alpha(h)} = \frac{\bar{\alpha}}{1+e^{-4h/\bar{h}}}$. The probability distributions assumed for $\bar{\boldsymbol{\alpha}}$ and $\bar{\boldsymbol{h}}$ are listed in Table S6.

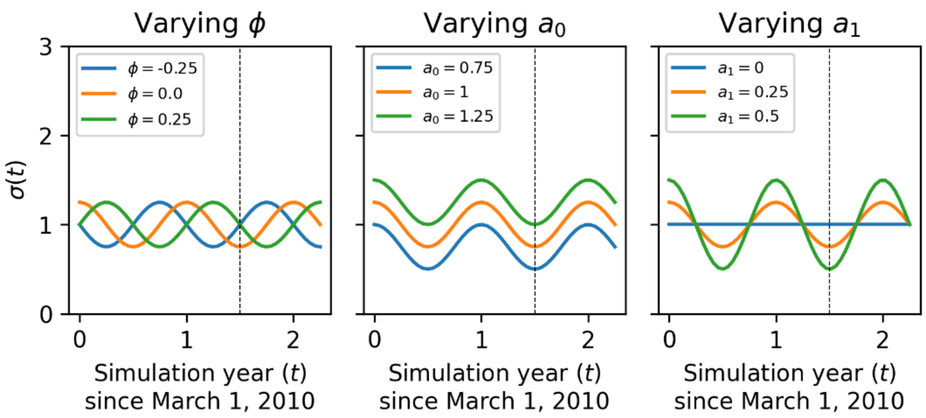


**Fig. S4: Modeling seasonality effect according to the function $\boldsymbol{\beta(t) = a_0 + a_1 \cos 2\pi(t - \phi)}$.** The vertical dotted lines marks September 1, 2021.

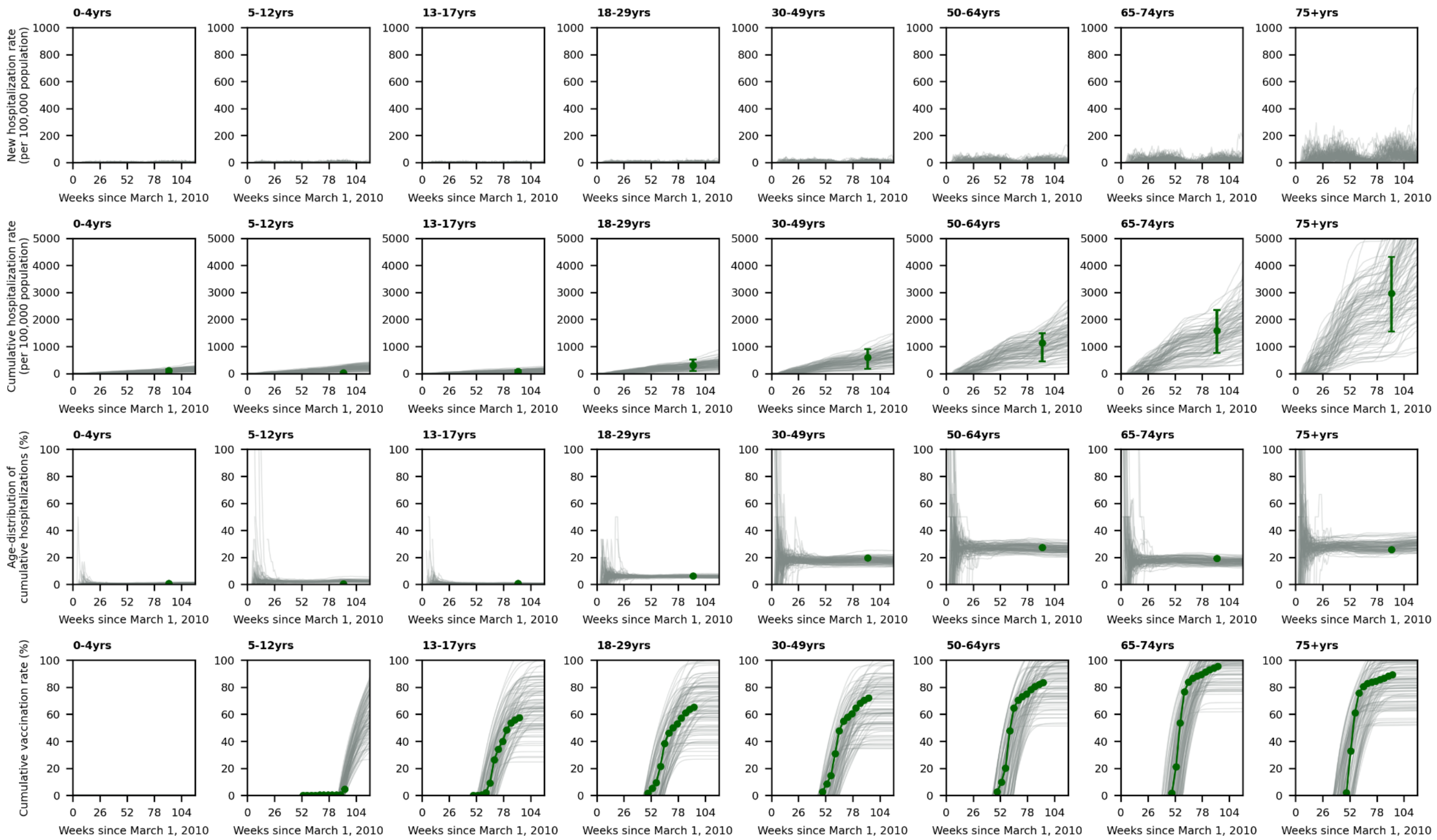


**Fig. S5: Rates of weekly hospitalizations and cumulative hospitalizations, age-distribution of cumulative hospitalizations, and cumulative vaccination rates by age.** The week 91 marks the beginning of winter 2022.

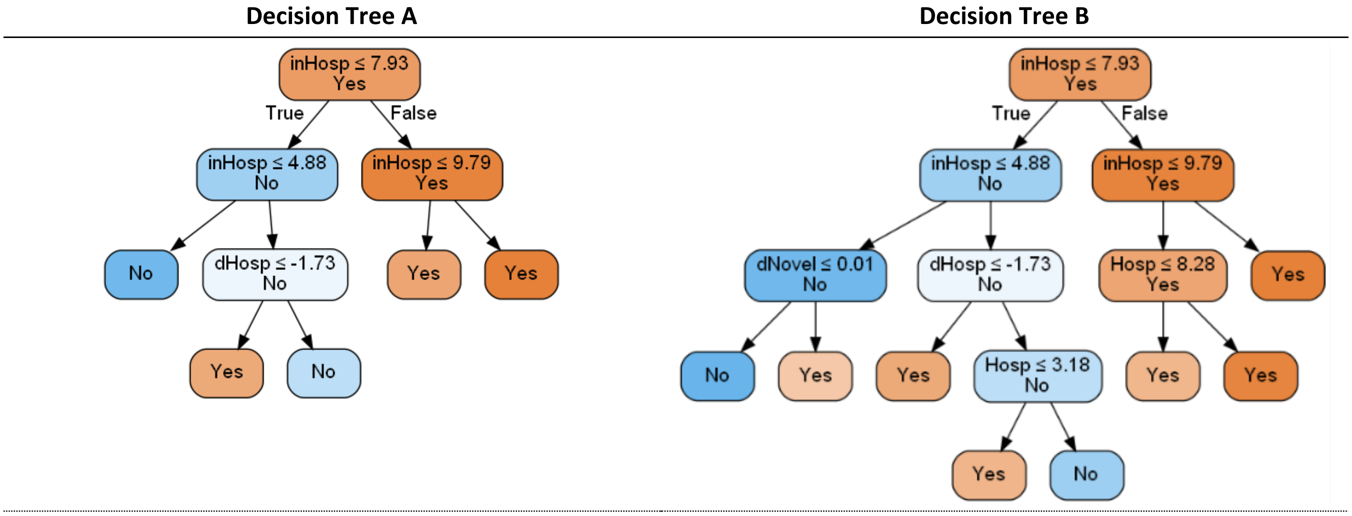


- **inHosp**: Current hospital occupancy due to COVID-19 (per 100,000 population).
- **Hosp**: Rate of weekly new COVID-19 hospitalizations averaged over the past 2 weeks (per 100,000 population).
- **dHosp**: Change in weekly new COVID-19 hospitalizations over the past 4 weeks (per 100,000 population).

**Fig. S6: Decision Trees A and B to predict whether the hospital occupancy due to COVID-19 would surpass the threshold of 10 per 100,000 population within the next 8 weeks during the winter and spring of 2022.**

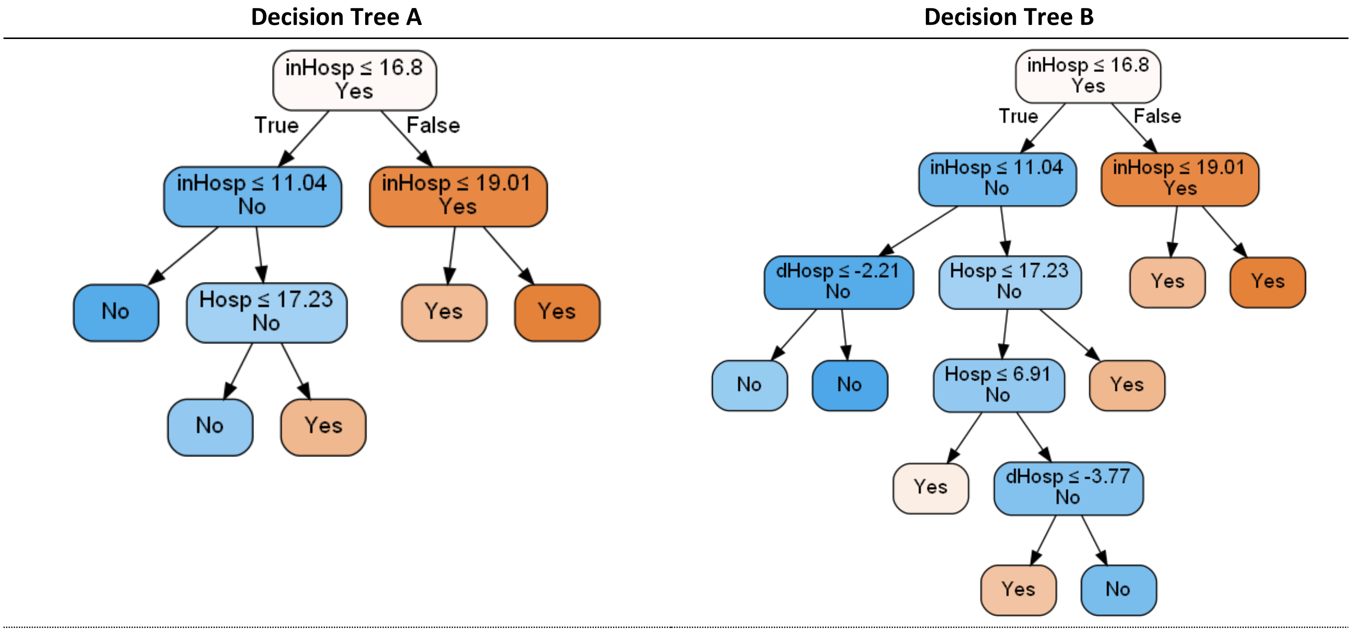


- **inHosp**: Current hospital occupancy due to COVID-19 (per 100,000 population).
- **Hosp**: Rate of weekly new COVID-19 hospitalizations averaged over the past 2 weeks (per 100,000 population).
- **dHosp**: Change in weekly new COVID-19 hospitalizations over the past 4 weeks (per 100,000 population).

**Fig. S7: Decision Trees A and B to predict whether the hospital occupancy due to COVID-19 would surpass the threshold of 20 per 100,000 population within the next 8 weeks during the winter and spring of 2022.**

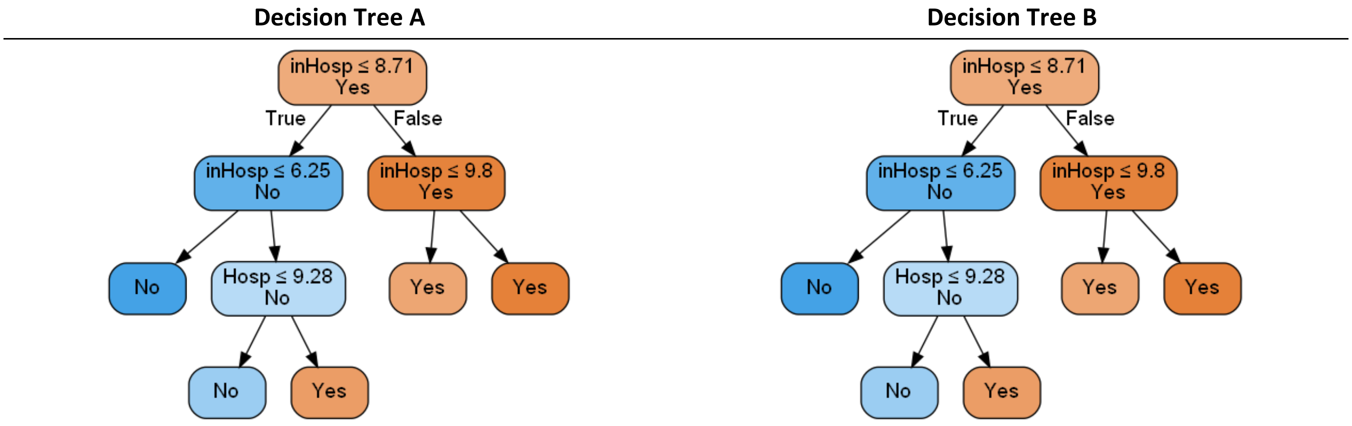


- **inHosp**: Current hospital occupancy due to COVID-19 (per 100,000 population).
- **Hosp**: Rate of weekly new COVID-19 hospitalizations averaged over the past 2 weeks (per 100,000 population).
- **dHosp**: Change in weekly new COVID-19 hospitalizations over the past 4 weeks (per 100,000 population).

**Fig. S8: Decision Trees A and B to predict whether the hospital occupancy due to COVID-19 would surpass the threshold of 10 per 100,000 population within the next 4 weeks during the winter and spring of 2022.**

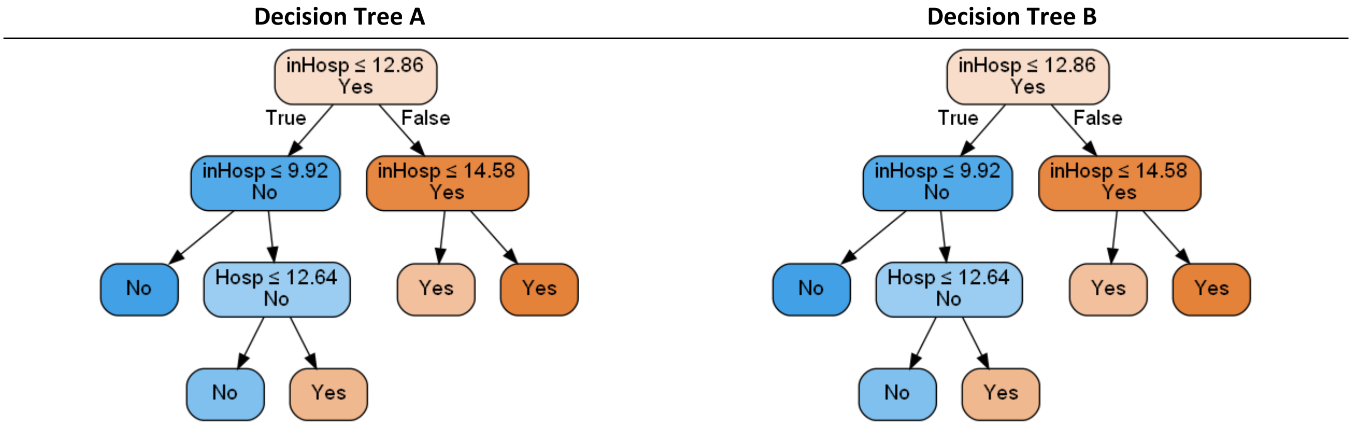


- **inHosp**: Current hospital occupancy due to COVID-19 (per 100,000 population).
- **Hosp**: Rate of weekly new COVID-19 hospitalizations averaged over the past 2 weeks (per 100,000 population).
- **dHosp**: Change in weekly new COVID-19 hospitalizations over the past 4 weeks (per 100,000 population).

**Fig. S9: Decision Trees A and B to predict whether the hospital occupancy due to COVID-19 would surpass the threshold of 15 per 100,000 population within the next 4 weeks during the winter and spring of 2022.**

**Decision Tree A** | **Decision Tree B**

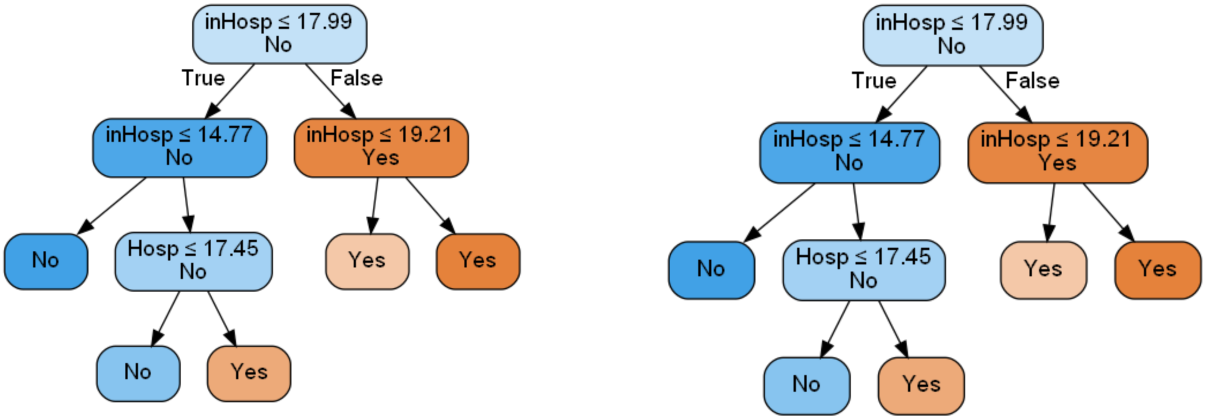


- **inHosp**: Current hospital occupancy due to COVID-19 (per 100,000 population).
- **Hosp**: Rate of weekly new COVID-19 hospitalizations averaged over the past 2 weeks (per 100,000 population).
- **dHosp**: Change in weekly new COVID-19 hospitalizations over the past 4 weeks (per 100,000 population).

**Fig. S10: Decision Trees A and B to predict whether the hospital occupancy due to COVID-19 would surpass the threshold of 20 per 100,000 population within the next 4 weeks during the winter and spring of 2022.**
